# Evidence for ApoE receptor 2-Disabled homolog-1 pathway disruption in the amygdala in sporadic Alzheimer’s disease

**DOI:** 10.1101/2025.06.13.25329511

**Authors:** Christopher E. Ramsden, Mark S. Horowitz, Daisy Zamora, Thomas G. Beach, Geidy E. Serrano, Richard A. Arce, Andrea Sedlock, Sophie Nagle, Rina Q. Shou, Fred E. Indig, John M. Davis, Dragan Maric

**Author notes:** Corresponding author: Christopher Ramsden, MD, PhD., Chief, Lipid Peroxidation Unit, Laboratory of Clinical Investigation, NIA/NIH, Baltimore, MD, 21224.

## Abstract

**INTRODUCTION:** The ApoE receptor 2-Disabled homolog-1 (ApoER2-Dab1) pathway suppresses Tau phosphorylation as part of a multi-arm pathway that regulates cytoskeletal and synaptic integrity. We previously showed that multiple ApoER2-Dab1 pathway components accumulate in regions affected in early Alzheimer’s disease (AD). Since the amygdala is a hub for emotional regulation and fear memory, we hypothesized that accumulation of ApoER2-Dab1 components in amygdala may correlate with cognitive or neuropsychiatric manifestations of AD.

**METHODS:** We used single-marker and multiplex immunohistochemistry to label ApoER2-Dab1 components in amygdala from 32 cases spanning the clinicopathological spectrum of AD.

**RESULTS:** Seven ApoER2-Dab1 pathway components accumulated in amygdala and correlated with histological progression and cognitive or neurobehavioral deficits in AD. ApoER2-Dab1 components accumulated within ApoER2-expressing neurons and dystrophic neurites surrounding ApoE-enriched extracellular plaques.

**DISCUSSION:** Findings add to growing evidence implicating ApoER2-Dab1 disruption in neurodegeneration and suggest that ApoER2-Dab1 disruption in amygdala may contribute to neuropsychiatric manifestations of AD.

## 1 Introduction

Neuropsychiatric symptoms—including anxiety, agitation, aggression, and depression [1]—are common manifestations of sporadic Alzheimer’s disease (AD) [2, 3] that are associated with more rapid progression to severe dementia and death.[4] Anxiety and depression contribute to personal suffering of AD patients, while agitation and aggression can increase caregiver stress and the need for 24-hour care and institutionalization,[5] thus adding to family and societal costs. As the major processing center for emotions,[6] the amygdala plays a central role in the regulation of mood, anxiety, agitation, and aggression.[7] The amygdala also plays a prominent role in connecting emotions to memory [8–10] and in the processing and recall of fear memories.[11] The amygdala is known to degenerate early in some patients with mild cognitive impairment (MCI) and AD.[12–16] The ApoE protein is strongly enriched in the detergent-insoluble proteome of amygdala in dementia cases [17] and the *APOE* ε4 allele has recently been linked to increased aggression and agitation in AD,[18, 19] implying that altered brain lipid and lipoprotein metabolism in the amygdala could contribute to neuropsychiatric aspects of AD. However, the underlying mechanisms and neuropsychiatric manifestations of amygdala degeneration in AD are incompletely understood.

### 1.1 Does ApoER2-Dab1 pathway disruption underlie amygdala neurodegeneration in AD?

The ApoE receptor 2-Disabled homolog-1 (ApoER2-Dab1) pathway is a three ligand, multi-arm signaling cascade that regulates cytoskeletal and synaptic integrity and internalization of lipoparticles [20–25] (reviewed in [25–27]). Activation of the ApoER2-Dab1 pathway suppresses Tau phosphorylation as part of a four-arm pathway that regulates stability of the microtubule cytoskeleton (via Tau phosphorylation), the actin cytoskeleton (via LIMK1 phosphorylation), synapse strength (via PSD95 phosphorylation), and neuronal delivery of cholesterol and specialized phospholipids (via lipoprotein internalization) (reviewed in [25, 27]). Disruption of this pathway at the level of ApoER2 could potentially trigger four core molecular derangements implicated in AD pathogenesis, while inducing co-accumulation of multiple pathway components.[25] We previously showed that multiple ApoER2-Dab1 pathway components accumulate together with hyperphosphorylated Tau (pTau) in five regions known to degenerate in the earliest stages of AD (entorhinal cortex (ErC), prosubiculum-CA1 border region, temporal neocortex, locus coeruleus, raphe nucleus),[25] as well as two regions that degenerate later in the disease process (molecular layer of dentate gyrus and hippocampus).[27] In each of these seven regions, accumulation of both extracellular ApoER2 ligands and multiple neuronal ApoER2-Dab1 signaling partners correlated with clinicopathological progression of AD. Collective findings suggested that pTau accumulation may be only one of many consequences stemming from ApoER2-Dab1 pathway disruption, and formed the basis for a unifying AD model that integrates pTau lesions with other hallmark (ApoE, amyloid β (Aβ), ApoJ) and emerging (Reelin, Dab1, pP85α_Tyr607_, pPSD95_Thr19_) neuropathological features.

However, it is not yet known if ApoER2-Dab1 pathway disruption plays a role in amygdala degeneration in MCI and AD, or whether accumulation of ApoER2 pathway components correlates with cognitive or neuropsychiatric endpoints.

We therefore used immunohistochemistry (IHC) to search for evidence of ApoER2-Dab1 pathway disruption in amygdala specimens from 32 rapidly autopsied individuals who died cognitively normal, with MCI, or with AD dementia. We found that: (1) ApoER2 is highly expressed by a subset or amygdala neurons; (2) ApoER2 accumulates together with five of its neuronal signaling partners (Dab1, pP85α_Tyr607_, pLIMK1_Thr508_, pTau _Ser202/Thr205_ and pPSD95_Thr19_) and one of its extracellular ligands (ApoJ) in abnormal neurons/neurites and extracellular plaques, respectively, in MCI and AD cases; and (3) accumulations of ApoER2-Dab1 pathway components correlated with histological progression, cognitive deficits, and neuropsychiatric endpoints. Multiplex-IHC revealed that pLIMK1_Thr508_, pTau _Ser202/Thr205_ and pPSD95_Thr19_ accumulate together within many of the same abnormal neurons and dystrophic dendrites in the vicinity of ApoE- and Aβ-enriched extracellular plaques, while Dab1 accumulated in both MAP2-labeled dendrites and NFL-labeled dystrophic axons surrounding ApoE-Aβ plaques.

Findings suggest that disruption of this pathway in amygdala may contribute to synapse dysfunction and the cognitive and neuropsychiatric manifestations of AD.

## 2 Materials and Methods

### 2.1 Case selection and postmortem specimens

We obtained formalin-fixed, paraffin-embedded (FFPE), six micron-thick coronal amygdala sections from 32 rapidly autopsied cases spanning the clinicopathological spectrum of AD (**Supplementary Tables 1-2**) from Arizona Study of Aging and Neurodegenerative Disorders and Brain and Body Donation Program (BBDP) at the Banner Sun Health Research Institute (http://www.brainandbodydonationprogram.org)[28]. To mitigate limitations due to tissue degradation, BBDP employed a rapid on-call autopsy team to achieve short postmortem interval (PMI) (mean 3 h). Standardized minimal fixation procedures were employed using 1 cm^3^ tissue blocks fixed in 10% neutral buffered formalin for 48 hrs. BBDP subjects provided written consent for study procedures, autopsy and sharing of de-identified data prior to enrollment. The study and its consenting procedures were approved by the Western-Copernicus Group

Institutional Review Board (IRB) of Puyallup, Washington, and was conducted in accordance with the ethical standards as laid down in the 1964 Declaration of Helsinki. The BBDP population has been extensively described.[28–31] Most donors were enrolled as cognitively normal volunteers residing in retirement communities near Phoenix, Arizona, USA. Following informed consent, donors received standardized medical, neurological, and neuropsychological assessments. Neuropathological and cognitive endpoints captured by BBDP are described in previous publications [28–31] and in the supplementary methods. As previously described,[25, 27] AD cases selected for this study had clinical dementia during life with pathologic diagnosis determined according to the NIA-Reagan criteria [32] using a high likelihood of AD threshold. MCI cases selected for this study were classified by BBDP based on a clinical diagnosis plus the presence of mild-to-moderate AD-type pathology that did not meet NIA-Reagan criteria for AD. Controls did not meet criteria for AD or MCI, however some degree of AD-type pathology was evident at autopsy in most controls. Key individual and summary characteristics of the BBDP cohort including PMI, Braak stage (0-VI), Thal phase (0-5), total amyloid plaques (0-15), neuritic plaque density (0-3), and *APOE* status are provided in **Supplementary Tables 1-2**. The antemortem Mini-Mental Status Exam (MMSE, 0-30) test had the least missing data and was selected as the main cognitive endpoint. A detailed description of the National Alzheimer’s Coordinating Center (NACC) Uniform Data Set is provided in **Supplementary Table 3**.

Exploratory analyses used four neuropsychological endpoints from the NACC Uniform Data Set—behavior, comportment, and personality (Form B4, Item 9), personality change (Form B9, Item 9g), behavioral symptoms (Form B9, Item 8), and depression (Form B6, Item 16; also known as the Geriatric Depression Scale-15 (GDS-15)).

### 2.2 Single-marker immunohistochemistry (IHC) and antibody validation

Single-marker IHC was performed by Histoserv (Gaithersburg, MD, USA) and in our NIA laboratory, as previously described.[25, 27] Combined blocks containing cortex from AD cases and non-AD controls were sectioned into 6μm-thick coronal sections. Optimal immunostaining conditions were then empirically determined using an incremental heat induced epitope retrieval (HIER) method using 10 mM Na/Citrate pH6 and/or 10 mM TRIS/EDTA pH9 buffer heated at 70°C for 10-40min or formic acid (88%, 10-20 min) at room temperature.

Secondary antibodies (Jackson ImmunoResearch) used for single IHC chromogenic staining were matched to the host class/subclass of the primary antibody. These combined AD plus anatomically-matched tissue blocks were used to generate negative controls (comparing staining results versus pre-immune serum and with the primary antibody omitted) and positive controls (comparing staining results in temporal or frontal cortex specimens from confirmed AD cases with known region-specific Aβ and tangle scores versus non-AD controls).[25, 27] Empirically determined optimal conditions were then used to immunolabel sets of slides using IHC and MP-IHC. Briefly, for IHC, sections were first deparaffinized using standard Xylene/Ethanol/Rehydration protocol followed by antigen unmasking with 10–40 min HIER at 70°C or formic acid for 10-20 min at room temperature. Sources and technical specifications of reagents used in this study are provided in **Supplementary Table 4**. All antibodies used for IHC in this study have previously been used for IHC in human FFPE specimens and those targeting core ApoER2-Dab1 pathway components have been used specifically for IHC in human brain specimens in published manuscripts.[25, 27]

### 2.3 Multiplex fluorescence immunohistochemistry, image acquisition and computational reconstruction

MP-IHC was performed as previously described [25, 27] to provide cytoarchitectural, spatial and pathological context for single-marker IHC images, which were used for quantitation. Briefly, coronal amygdala sections were mounted on Leica Apex Superior adhesive slides (VWR, 10015-146) to prevent tissue loss. Six iterative rounds of sequential MP-IHC staining were completed with antibody panels targeting the ApoER2-Dab1 pathway, classical AD biomarkers, and cytoarchitectural biomarkers to map brain tissue parenchyma (see **Supplementary Table 4**). Fluorophore-labelled protein targets from each round of staining were imaged by multispectral epifluorescence microscopy followed by antibody stripping and tissue antibody re-staining steps to repeat the cycle,[25, 27, 33] each time using a different antibody panel. Image tiles (600×600μm viewing area) were individually captured using a 10-color Zeiss AxioImager.Z2 epifluorescence microscope at 0.325 micron/pixel spatial resolution, as previously described,[33] and the tiles stitched into whole specimen images using the ZEN 2 image acquisition and analysis software program (Zeiss), with an appropriate color table having been applied to each image channel to either match its emission spectrum or to set a distinguishing color balance. The RGB histogram of each image was adjusted using Zen software, resulting in optimized signal brightness and contrast, improved dynamic range, exposure correction, and gamma/luminosity-enhancement to reveal hidden/dim image details.

Stitched images were exported as BigTIFF files, then computationally registered at the subpixel level using affine transformation and corrected for photobleaching, autofluorescence, non-uniform illumination shading, spectral bleed-through, and molecular colocalization artifacts, as previously described.[33] Images were exported as BigTIFF and imported into Adobe large document format upon which the images were linearly contrast-enhanced using the levels function, sharpened to reduce blurring using the sharpening filter, and pseudo-colored to enhance color contrast either to show colocalization or separated to display multiple markers in a single image, as previously described.[25, 27, 34, 35]

### 2.4 Regional Annotation and Quantitation

Single-marker IHC images were analyzed using HALO 3.5 (Indica Labs, Corrales, NM). The amygdala and ErC regions were identified using a combination of anatomical landmarks and cytoarchitectonic mapping as previously described [25, 27] with additional confirmation by Banner neuropathologist (GS). No attempt was made to identify individual amygdala nuclei. If the cytoarchitecture was distinct, the flood fill annotation tool was used to help define boundaries and to limit the inclusion of edge artefacts. Otherwise, the brush annotation tool was used.

Following the registration of serial slides, regional annotations were copied to each image. For most pathological markers, HALO’s Area Quantification v2.4.2 module was used to quantify the stain positive area as a percentage of each annotated region, excluding any colocalized cell nuclei.[25, 27] Although prominent accumulations of Dab1 were evident in dystrophic neurites in the immediate vicinity of neuritic plaques, it is also expressed by many healthy neurons. To distinguish between pathological plaque-associated Dab1 and the Dab1 typically present in neurons, we used HALO’s Object Colocalization v2.1.5 module, with an embedded classifier to narrow the analysis area, to quantify plaque-associated Dab1 objects per mm^2^.[25, 27]

### 2.5 Statistical analysis

For each annotated region, between-group differences according to each marker were quantified using Kruskal-Wallis tests. Variable transformations (e.g., natural log) were used as necessary. A Spearman’s correlation coefficient between each immunohistochemical marker and each AD endpoint (Braak stage, Aβ plaque load, cerebral amyloid angiopathy (CAA) score, MMSE) and neuropsychological endpoint (comportment, behavioral symptoms, personality change, depression) was calculated. Graphs showing individual data points in each group and their relationships to AD endpoints are provided in **Figs 1-5** and **7**. In a sensitivity analysis accounting for the false discovery rate, we adjusted the *P* values using the two-stage linear step-up procedure described by Benjamini et al. and Anderson.[36, 37] (**Supplementary Table 5**). Statistical analyses were conducted in Stata Release 19.[38]

**Figure 1.**
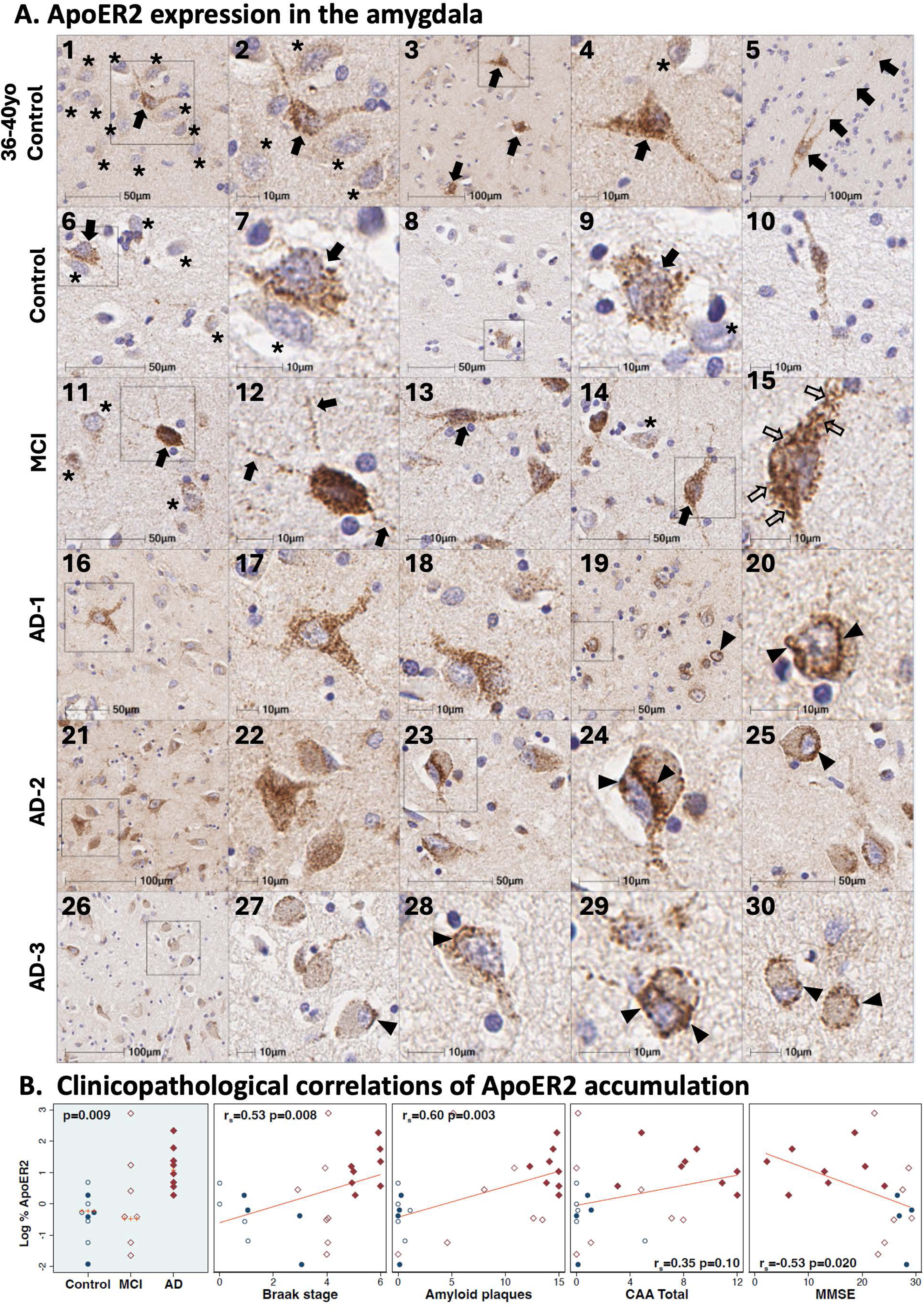
ApoER2 expression in the amygdala in controls, MCI and Alzheimer’s disease. Panel **A** includes single-target IHC images from coronal sections of the amygdala from a middle-aged Braak stage 0 control (**A_1-5_**), age-matched Braak stage I control (**A_6-10_**), Braak stage III MCI case (**A_11-15_**) and three AD cases: Braak stage VI (**A_16-20_**), Braak stage VI (**A_21-25_**), and Braak stage V (**A_26-30_**). In non-AD controls, ApoER2 is strongly expressed in soma and neuritic projections of a small subset of pyramid-shaped neurons (depicted by arrows in **A_1-4_**), with minimal or absent expression in many neighboring neurons (depicted by * in **A_1-4_**). The zoomed in image in panel **A_2_** shows four neighboring neurons with minimal or absent ApoER2 expression (depicted by *). Moderate-to-strong ApoER2 expression was also observed in rare, large polymorphic neurons with long (>100 μm) neuritic extensions, residing in white matter tracts adjacent to amygdala neuron clusters (arrows in **A_5_**). In controls without AD or MCI, ApoER2 exhibited a granular, homogenous pattern of expression throughout the soma and neurites (**A_2,_ A_4,_ A_7,_ A_9_**). By contrast, most neurons in AD and MCI cases exhibited prominent, enlarged ApoER2-labeled vacuolar structures (depicted by arrows in **A_15,_ A_20,_ A_24_**). Peri-nuclear ApoER2 accumulations were observed exclusively in AD cases and were most prominent in neurons with morphological abnormalities consistent with neurodegeneration (depicted by arrowheads in **A_19-20,_ A_24-25_**, and **A_27-30_**). ApoER2 expression increased across the clinicopathological spectrum of AD (**B**) and positively correlated with Braak stage, total amyloid plaques, and antemortem cognitive deficits, but not CAA score. Open and closed blue circles in Panel B indicate middle-aged controls and age-matched controls; open and closed red diamonds indicate MCI cases and AD cases, respectively.

## 3 Results

Using a combination of *in situ* hybridization, single-target IHC and multiplex fluorescence IHC in seven vulnerable brain regions, we previously showed that: (1) the ApoER2 (protein) and *LRP8* (gene) are strongly expressed in the same regions, layers, and neuron populations that develop pTau pathology in the earliest stages of AD;[25] (2) in MCI and AD, pTau is only one of many neuronal ApoER2 signaling partners that accumulate together within abnormal neurons;[25, 27] and (3) these ApoER2 signaling partners accumulate in dystrophic neurites that surround extracellular ApoER2 ligands, including ApoE, ApoJ, and Reelin (i.e. neuritic plaques).[25, 27] Here, in amygdala we sought to characterize the distribution of ApoER2 expression and to determine whether neuronal ApoER2 signaling partners and extracellular ApoER2 ligands accumulate in abnormal neurons and neuritic plaques, respectively.

### 3.1 ApoER2 expression in the human amygdala

Single-target IHC in non-AD controls revealed that ApoER2 protein is strongly expressed in the soma and neuritic projections of a small subset of pyramid-shaped amygdala neurons (depicted by arrows in **Fig 1A_1-4_**); expression was minimal or absent in many neighboring neurons (depicted by * in **Fig 1A_1-4_**). This non-homogenous ApoER2 expression pattern in amygdala differs from the highly laminar ApoER2 expression pattern that we previously observed in human ErC and hippocampal formation.[25, 27] Moderate-to-strong ApoER2 expression was also evident in rare, large polymorphic neurons residing in white matter tracts adjacent to neuron clusters (arrows in **Fig 1A_5_** label a large polymorphic neuron with long (>100um) neuritic extensions. In controls without AD or MCI, ApoER2 exhibited a granular and relatively homogenous pattern of expression throughout the soma and neurites (**Fig 1A_2_**, **A_4_**, **A_7_**, **A_9_**). By contrast, many (but not all) neurons in AD and MCI cases exhibited prominent, enlarged ApoER2-labeled vacuolar structures (arrows in **Fig 1A_15_**, **A_20_**, **A_24_**). Peri-nuclear ApoER2 accumulations were observed exclusively in AD cases and were most prominent in neurons with morphological abnormalities consistent with neurodegeneration (depicted by arrowheads in **Fig 1A_19-20,_ A_24-25_**, **A_27-30_**). ApoER2 expression increased across the clinicopathological spectrum of AD and positively correlated with Braak stage, total amyloid plaques, and antemortem cognitive deficits (**Fig 1B**), but not CAA score.

### 3.2 Pathological accumulation of ApoER2-Dab1 pathway components in the amygdala

**3.2.1 Neuronal accumulation of ApoER2 signaling partners in the amygdala in AD**

Single-target IHC revealed that five neuronal ApoER2 signaling partners (Dab1, pP85α_Tyr607_, pLIMK1_Thr508_, pTau_Ser202/Thr205_, pPSD95_Thr19_) accumulated in MCI and AD cases and positively correlated with histological progression and antemortem cognitive deficits (**Figs 2-3**). ApoER2 signaling partners exhibited different distributions and morphologies. Dab1 accumulated within globular structures consistent with plaque-associated dystrophic neurites and to a lesser extent within neuronal soma (**Fig 2, A_1-5_**). pP85α_Tyr607_ and pLIMK1_Thr508_ accumulated in neuronal vacuolar structures and a smaller subset of plaque-associated dystrophic neurites (**Fig 2, A_6-15_**). pTau prominently accumulated as hallmark neuropil threads and neurofibrillary tangles (NFTs), and in the neuritic components of neuritic plaques (**Fig 3, A_1-5_**). pPSD95_Thr19_ accumulated in vacuolar structures within neuronal soma, small punctae surrounding affected neurons, and globular structures consistent with plaque-associated dystrophic neurites (**Fig 3, A_6-10_**). The expression of Dab1, pP85α_Tyr607_, pLIMK1_Thr508_, pTau_Ser202/Thr205_, and pPSD95_Thr19_ increased across the clinicopathological spectrum of AD and positively correlated with Braak stage, Aβ plaque load, CAA score, and antemortem cognitive deficits (**Fig 2B, 3B**), with particularly strong associations observed for pP85α (**Fig 2B**), pTau (**Fig 3B**), and pPSD95 (**Fig 3B**).

**Figure 2.**
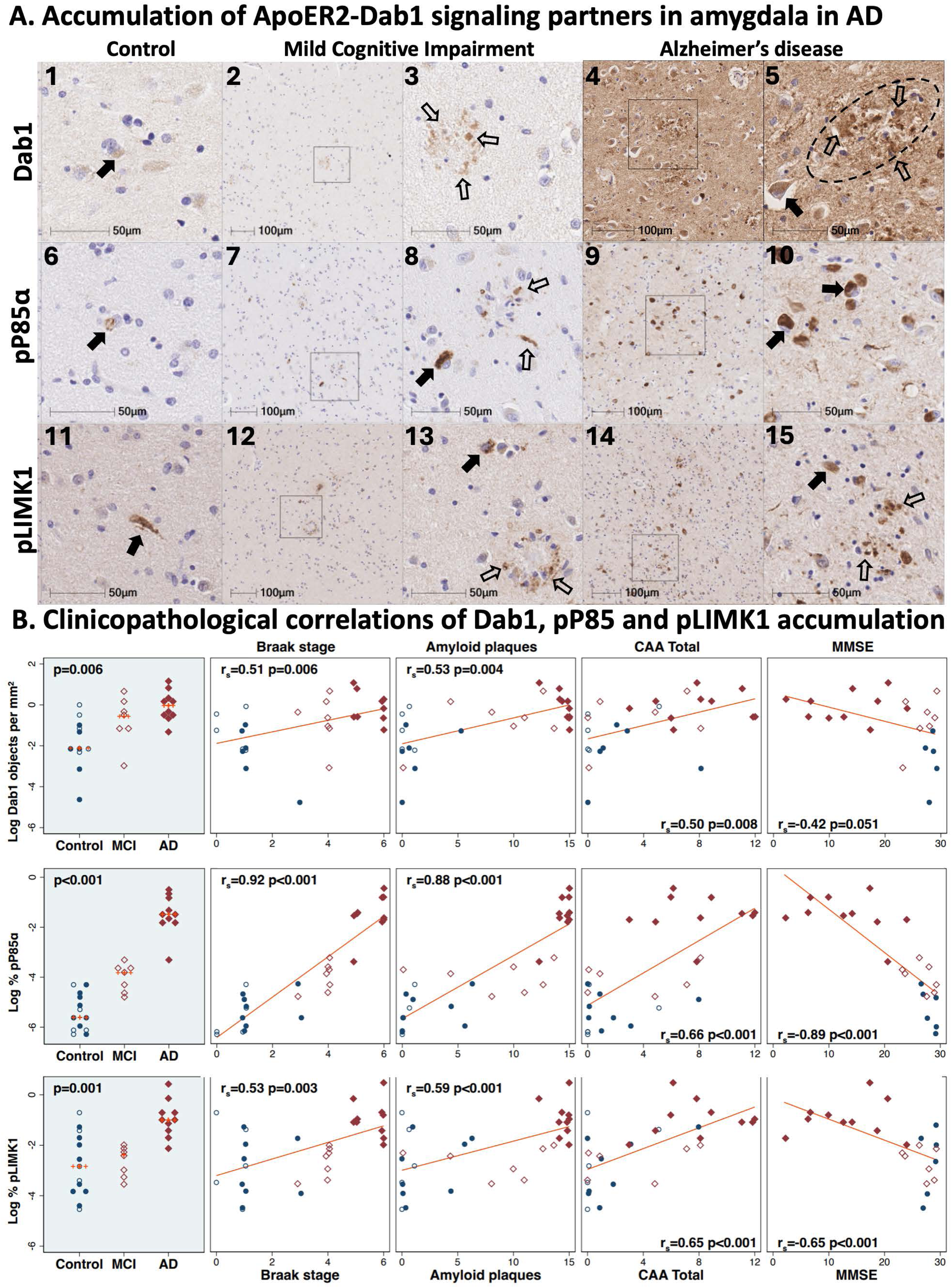
Neuronal accumulation of Dab1, pP85a, and pLIMK1 in the amygdala in MCI and AD cases. Serial coronal sections of the amygdala from a representative non-AD control (Braak stage III), MCI case (Braak stage IV), and AD case (Braak stage VI) were stained with anti-Dab1 (**A_1-5_**), anti-pP85α (**A_6-10_**), and anti-pLIMK1 (**A_11-15_**) antibodies. In AD cases, prominent accumulations of Dab1, pP85α_Tyr607_, and pLIMK1_Thr508_ were observed in abnormal neurons (designated by closed arrows in **A_5,_ A_10,_** and **A_15_**) and dystrophic neurites (designated by open arrows in **A_5_** and **A_15_**). Less pronounced accumulations were evident in MCI cases **A_2-3,7-8,12-13_**), with only sparse expression in non-AD controls. Dab1, pP85α_Tyr607_, and pLIMK1_Thr508_ expression increased across the clinicopathological spectrum of AD and positively correlated with Braak stage, Aβ plaque load, CAA score, and antemortem cognitive deficits (**B**), with particularly strong associations observed for pP85α (**B**). Open and closed blue circles in Panel **B** indicate middle-aged controls and age-matched controls; open and closed red diamonds indicate MCI cases and AD cases, respectively.

**Figure 3.**
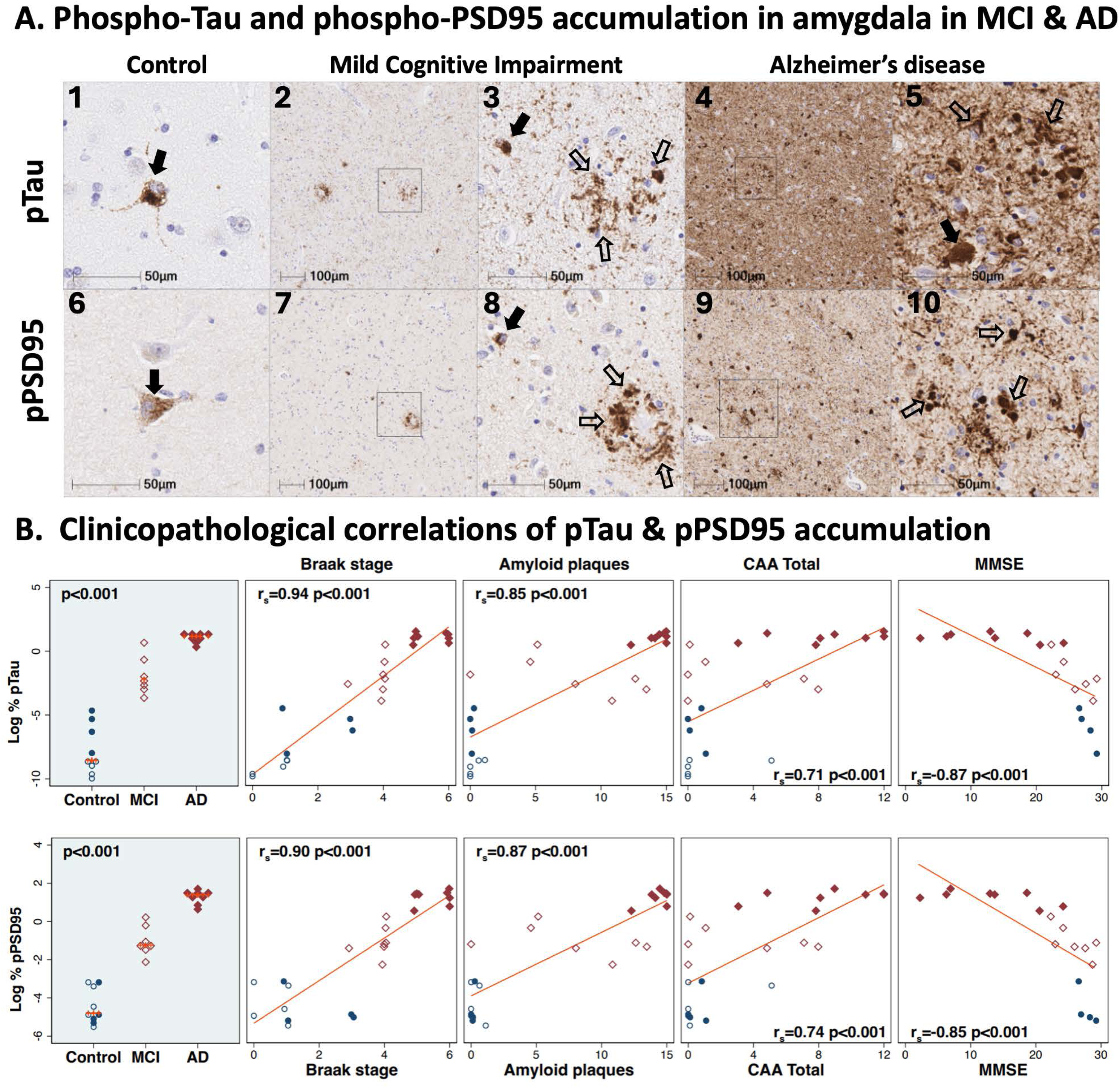
Phospho-Tau and phospho-PSD95 accumulate in the amygdala in MCI and AD. Serial coronal sections of the amygdala from a representative non-AD control (Braak stage III), MCI case (Braak stage IV), and AD case (Braak stage VI) were stained with anti-pTau (**A_1-5_**), and anti-pPSD95 (**A_6-10_**) antibodies. In MCI and AD cases, prominent accumulations of pTau and pPSD95 were observed in abnormal neurons (designated by closed arrows in **A_3,_ A_5_** and **A_8_**) and dystrophic neurites (designated by open arrows in **A_3,_ A_5,_ A_8_** and **A_10_**). Less pronounced accumulations were evident in MCI cases **A_2-3, 7-8, 12-13_**), with only sparse expression in non-AD controls. pTau and pPSD95 expression increased across the clinicopathological spectrum of AD and positively correlated with Braak stage, Aβ plaque load, CAA score, and antemortem cognitive deficits (**B**). Open and closed blue circles in Panel **B** indicate middle-aged controls and age-matched controls; open and closed red diamonds indicate MCI cases and AD cases, respectively.

#### 3.2.2 ApoER2 signaling partners and pTau accumulate together within abnormal neurons in AD

Having demonstrated regional co-accumulation of multiple ApoER2-Dab1 signaling partners in the amygdala in AD, we next sought to determine whether these pathway components accumulate together within the same pTau-expressing neurons, or within different cells. Single-target and multiplex-IHC amygdala images from a representative Braak stage V AD case (**Fig 4**) revealed that Dab1, pLIMK1_Thr508_, and pPSD95_Thr19_ accumulate together within many of the same pTau-expressing neurons and neurites (depicted by arrows in **Fig 4B_1-8_**). By contrast, little or no expression of Dab1, pLIMK1_Thr508_, or pPSD95_Thr19_ was observed in neighboring neurons that lacked pTau expression (depicted by * in **Fig 4B_1-8_**).

**Figure 4.**
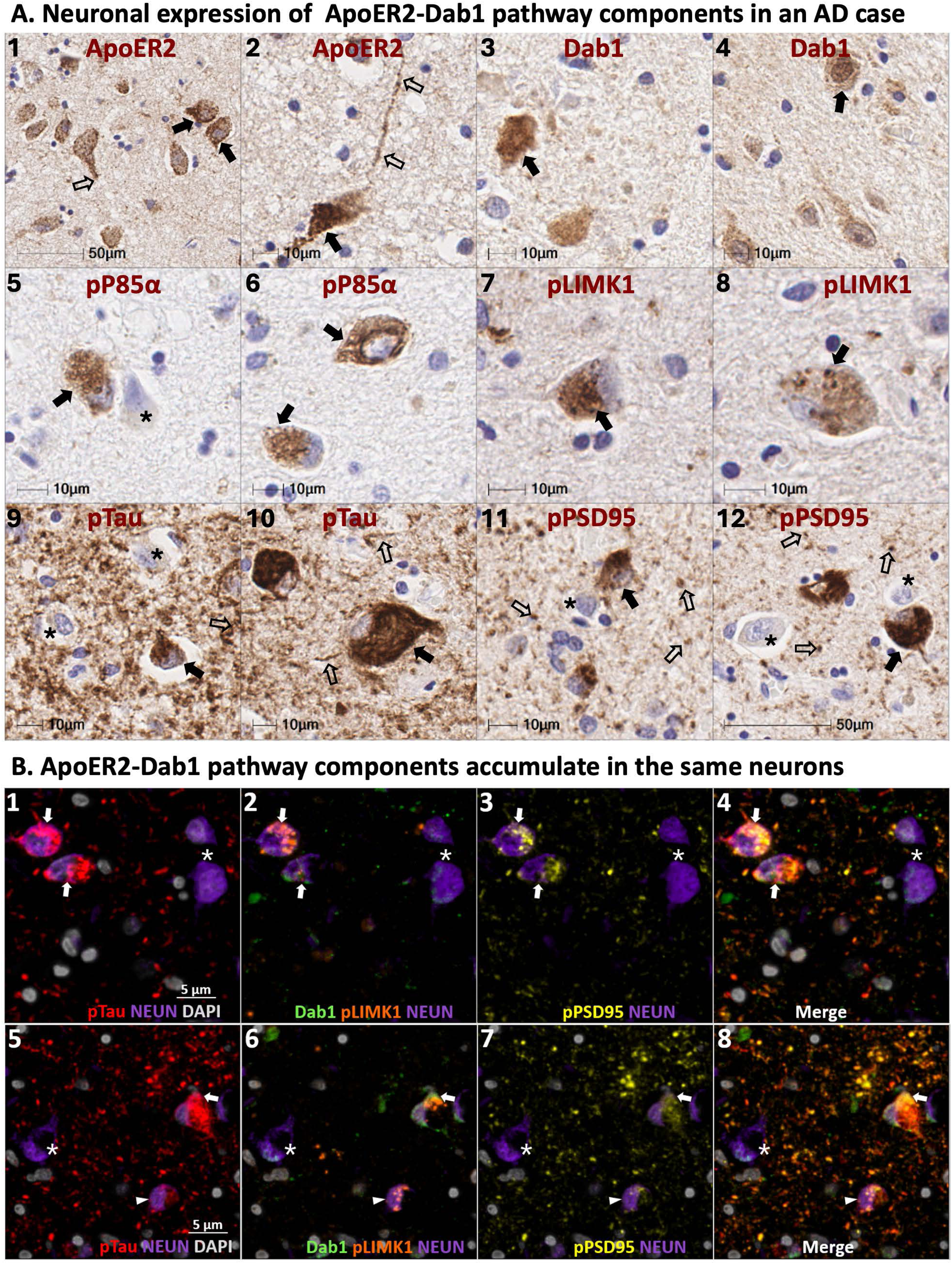
ApoER2-Dab1 pathway components co-accumulate in the same neurons in AD. Serial coronal sections of the amygdala from a representative AD case (Braak stage V) were stained with anti-ApoER2 (**A_1-2_**), anti-Dab1 (**A_3-4_**), anti-pP85α (**A_5-6_**), anti-pPLIMK1 (**A_7-8_**), anti-pTau (**A_9-10_**), and anti-pPSD95 (**A_11-12_**) antibodies. Single marker IHC demonstrates patterns for expression and accumulation of individual ApoER2-Dab1 pathway components within neuronal soma (designated by closed arrows in **A_1-12_**) and neurites (open arrows n **A_1-2_** and **A_9-12_**). Multiplex-IHC revealed that Dab1, pLIMK1_Thr508_, and pPSD95_Thr19_ accumulate together within many of the same pTau-expressing neurons (depicted by arrows in **B_1-8_**) and adjacent neurites. By contrast, little or no expression of Dab1, pLIMK1_Thr508_, or pPSD95_Thr19_ was evident in neighboring neurons that lacked pTau expression (depicted by * in **B_1-8_**). Arrowheads in **B_5-8_** depict one neuron with prominent granulovacuolar pLIMK1_Thr508_ expression, modest pPSD95_Thr19_ and pTau expression and with minimal Dab1 expression. Abbreviations: pTau, phosphorylated Tau; Dab1, disabled homolog-1; pLIMK1, Thr508-phosphorylated LIM kinase-1; pPSD95, Thr19-phosphorylated postsynaptic density 95; NEUN, neuron soma marker; DAPI, nuclear marker.

**Figure 5.**
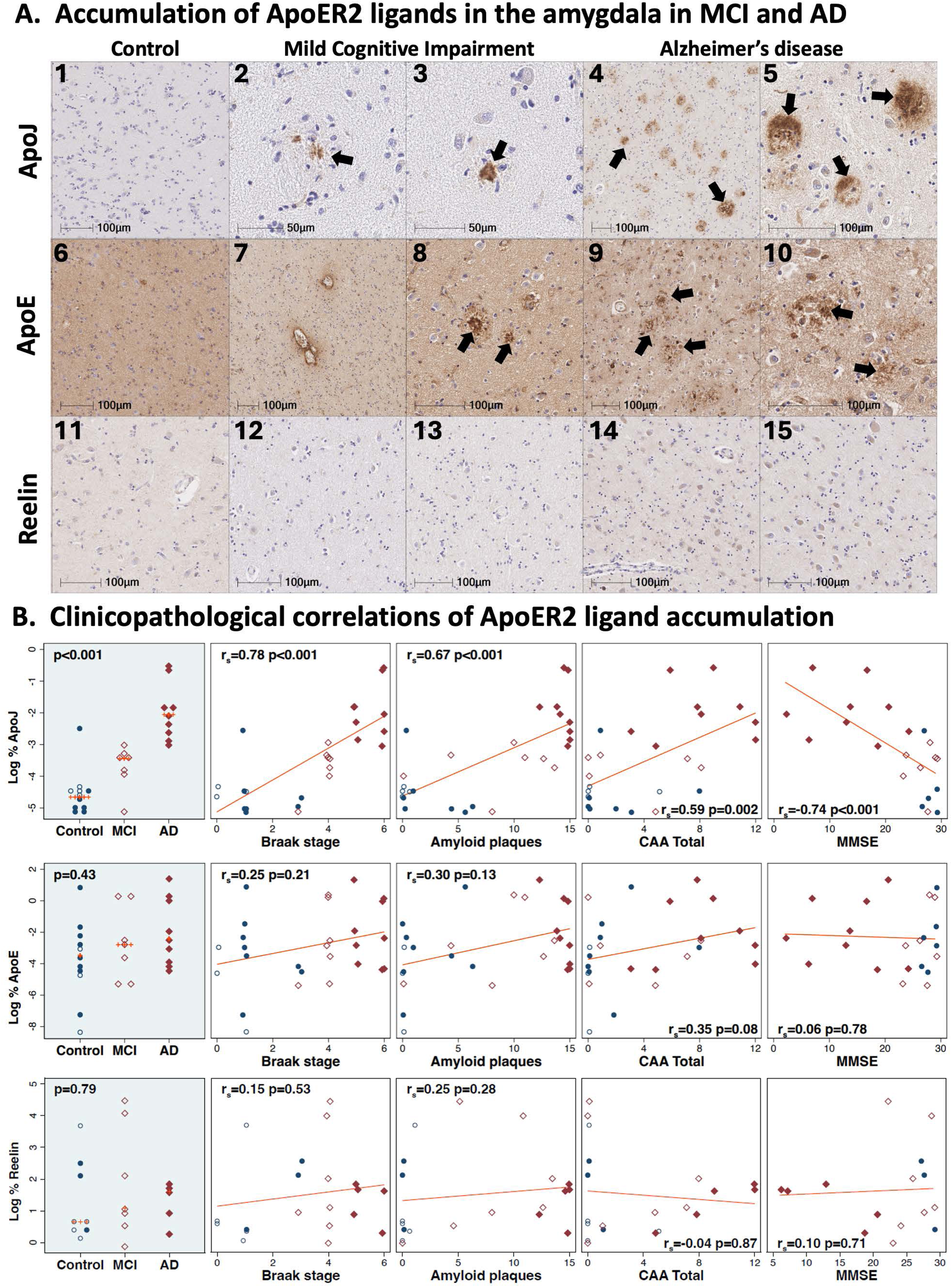
Accumulation of extracellular ApoER2 ligands in the amygdala in MCI and AD. Serial coronal sections of the amygdala from a representative non-AD control (Braak stage III), MCI case (Braak stage IV), and AD case (Braak stage VI) were stained with anti-ApoJ (**A_1-5_**), anti-ApoE (**A_6-10_**), and anti-Reelin (**A_11-15_**) antibodies. In AD cases, extracellular accumulations of ApoJ were frequent and prominent (designated by arrows in **A_4-5_**). Extracellular accumulations of ApoJ were sparse and less pronounced in MCI cases (arrows in **A_2-3_**), with only modest expression in non-AD controls (**A_1_**). ApoE accumulated within a subset of extracellular plaques and vascular lesions in AD and MCI cases (arrows in **A_8-10_**); however, substantial ApoE signals were observed in the parenchyma of all cases including non-AD controls. Minimal extracellular accumulation of Reelin was evident in amygdala (**A_11-15_**). ApoJ expression increased across the clinicopathological spectrum of AD and positively correlated with Braak stage, Aβ plaque load, CAA score, and antemortem cognitive deficits (**B**), Open and closed blue circles in Panel **B** indicate middle-aged controls and age-matched controls; open and closed red diamonds indicate MCI cases and AD cases, respectively.

#### 3.2.3 Extracellular accumulation of ApoER2 ligands in the amygdala in AD

Single-target IHC revealed prominent ApoJ accumulation within extracellular plaques (arrows in **Fig 5A_2-5_**), peri-vascular plaques, and vascular lesions including apparent CAA lesions in MCI and AD cases. ApoE similarly accumulated within a subset of extracellular plaques and vascular lesions (arrows in **Fig 5A_8-10_**). However, unlike ApoJ, substantial ApoE signals were observed in the parenchyma of controls and thus numerical differences in Controls, MCI and AD cases did not reach statistical significance (**Fig 5B**). Unlike ApoJ and ApoE, no extracellular accumulation of Reelin was observed in amygdala in MCI or AD (**Fig 5A_11-15_**). This lack of Reelin accumulation in amygdala differed from the prominent extracellular Reelin deposits that we previously observed in the hippocampus and subiculum in a subset of these same MCI and AD cases.[27] Since Reelin signaling through ApoER2 regulates the degradation and phosphorylation status of neuronal ApoER2 signaling partners including Dab1, P85α and Tau respectively in preclinical models, this extracellular Reelin deposition in hippocampus suggests that disruption of Reelin-ApoER2 binding and internalization in hippocampus may play a role in AD pathogenesis. By contrast, the lack of extracellular Reelin accumulation (**Fig 5A_11-15_**) in amygdala in the present study implies that Reelin depletion may play a more prominent role in amygdala.

#### 3.2.4 Co-accumulation of ApoER2-Dab1 pathway components in neuritic plaques in AD

Single-target stains using serial sections from the same representative Braak stage VI AD case demonstrated plaque-associated accumulations of ApoJ, ApoE, Dab1, pP85α_Tyr607_, pLIMK1_Thr508_, pTau_Ser202/Thr205_, and pPSD95_Thr19_ in the amygdala (**Fig 6A_1-12_**). However, the use of serial sections does not provide the spatial and cytoarchitectural context required to determine whether ApoER2-Dab1 pathway components accumulate together in the same neuritic plaques. Multiplex-IHC revealed that ApoE accumulates in the central core of many neuritic plaques, and that multiple ApoER2-Dab1 pathway components accumulate together in the immediate vicinity of these extracellular ApoE deposits (**Fig 6B_1-8_**). Consistent with recent findings in hippocampus,[27] pontine nuclei [25] and middle temporal gyrus,[25] we observed that Dab1 is enriched in MAP2-labeled dendrites, as well as NFL-labeled axons and synaptophysin-labeled pre-synaptic terminals (**Fig 6B_1-8_**). Tau accumulates predominantly in MAP2-labeled dendrites but is also present in NFL-labeled axon terminals (**Fig 6B_2_** and **B_8_**). By contrast, pPSD95 appears to be enriched exclusively in MAP2-labeled dystrophic dendrites and absent from NFL-labeled axons (**Fig 6B_2_** and **B_8_**). Multiplex also revealed close spatial relationships between ApoE, infiltrating microglia, and reactive astrocytes in the neuritic plaque niche (**Fig 6B_4_** and **B_8_**), consistent with known glia-mediated plaque clearance pathways.[39]

**Figure 6.**
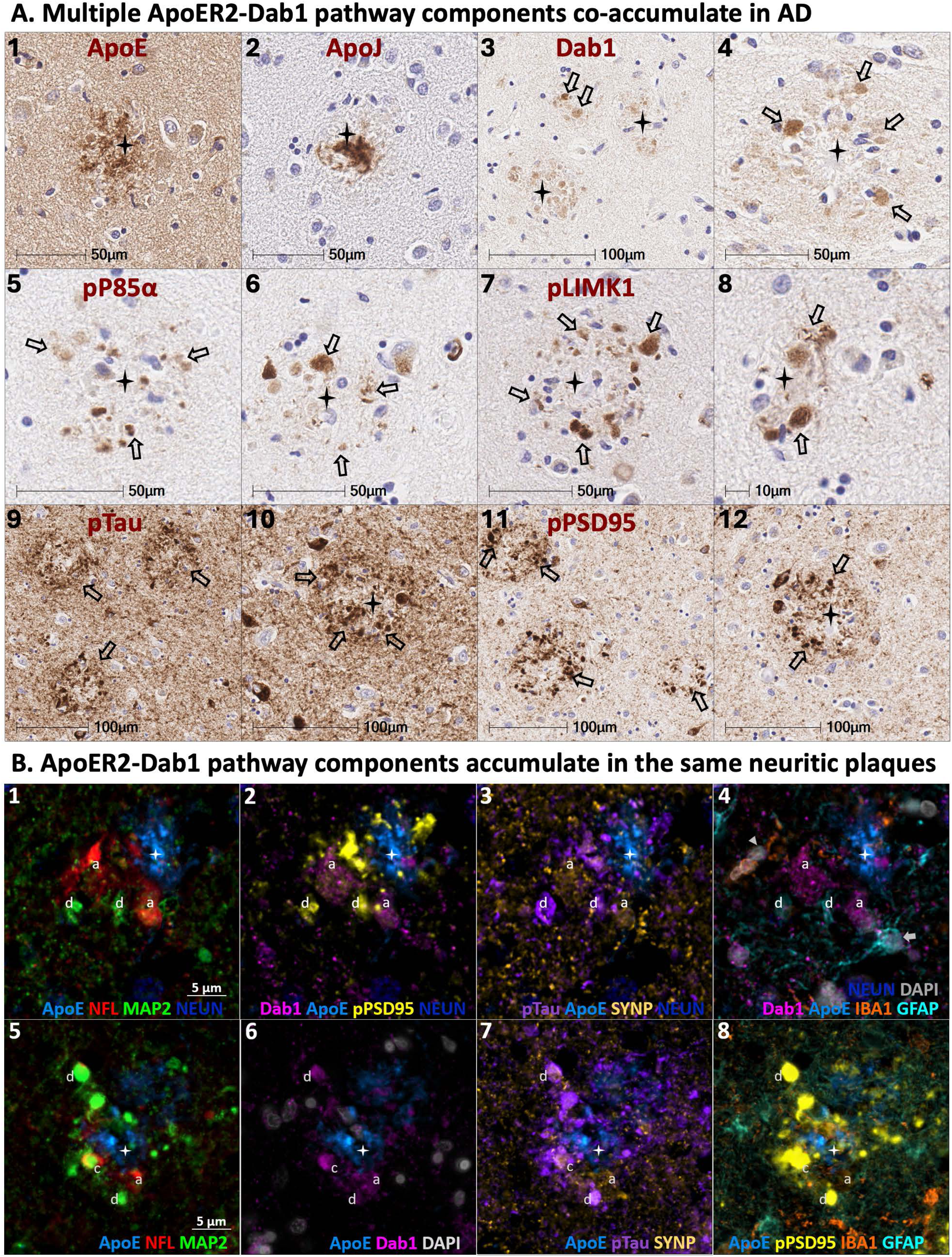
**ApoER2-Dab1 pathway components co-accumulate in neuritic plaques in AD** Serial coronal sections of the amygdala from a representative Braak stage VI AD case were stained with anti-ApoE (**A_1_**), anti-ApoJ (**A_2_**), anti-Dab1 (**A_3-4_**), anti-p85α (**A_5-6_**), anti-pPLIMK1 (**A_7-8_**), anti-pTau (**A_9-10_**), and anti-pPSD95 (**A_11-12_**) antibodies. Single marker IHC demonstrates patterns for expression and accumulation of individual ApoER2-Dab1 pathway components within extracellular plaques or adjacent dystrophic neurites (designated by stars and arrows, respectively in **A_1-12_**). Multiplex-IHC revealed that ApoE accumulates in the central core of many neuritic plaques, and that multiple ApoER2-Dab1 pathway components accumulate together in the immediate vicinity of these ApoE-enriched plaques (designated by stars in **B_1-8_**). Dab1 is enriched in both MAP2-labeled dendrites (designated by ‘d’ in **B_1-8_**) and NFL-labeled axons (‘a’ in **B_1-8_**). Tau accumulates predominantly in MAP2-labeled dendrites but is also present in NFL-labeled axon terminals while pPSD95 appears to be enriched in exclusively in MAP2-labeled dystrophic dendrites (**B_2-3_** and **B_7-8_**). Multiplex also revealed close spatial relationships between ApoE, IBA1-labeled infiltrating microglia and GFAP-labeled reactive astrocytes in the neuritic plaque niche (**B_4_** and **B_8_**). Abbreviations: ApoE, Apolipoprotein E; NFL, Neurofilament light chain; MAP2, Microtubule associated protein 2; NEUN, neuronal nuclear/soma antigen; Dab1, Disabled homolog-1; pPSD95, Thr19-phosphorylated postsynaptic density 95; pTau, phosphorylated Tau_Ser202/Thr205;_ SYNP, Synaptophysin; IBA1, ionized calcium-binding adapter molecule 1; GFAP, Glial fibrillary acidic protein; DAPI, nuclear marker.

**Figure 7.**
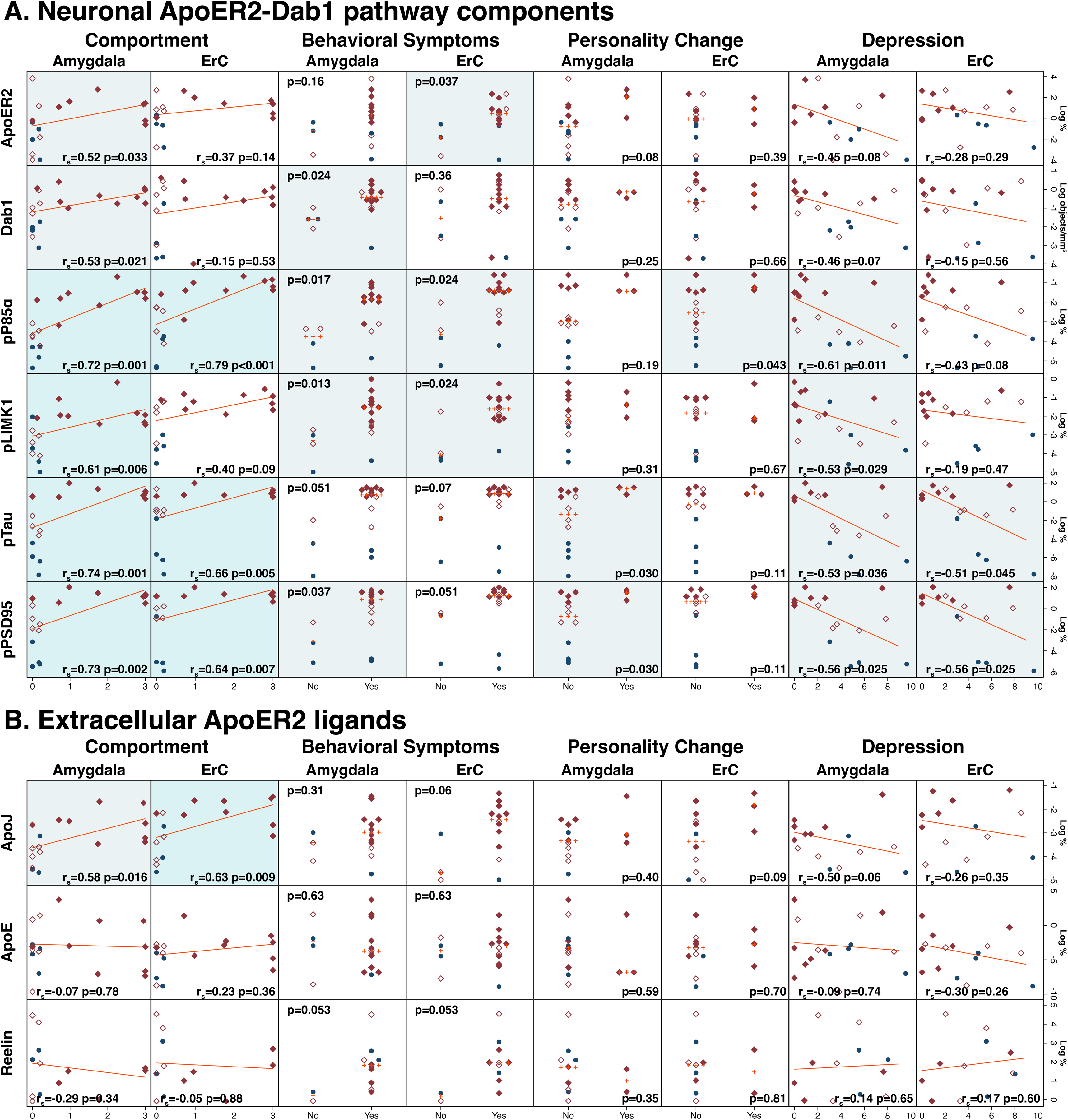
**Associations between ApoER2-Dab1 pathway components and neuropsychiatric endpoints** In amygdala, the expression levels of six neuronal ApoER2 signaling partners (**A**) and one extracellular ApoER2 ligand (ApoJ) (**B**) positively correlated with deficits in comportment. Several of these ApoER2-Dab1 pathway components also correlated with behavioral symptoms, personality changes, and depression. In ErC, expression levels of several ApoER2-Dab1 pathway components correlated with neuropsychiatric endpoints; however, these associations tended to be weaker and less robust. Dark, light blue, and white background indicate *P*<0.01, *P*<0.05, and *P*>0.05, respectively.

#### 3.2.5 ApoER2-Dab1 pathway components in amygdala correlate with neuropsychiatric endpoints in AD

We next sought to explore whether the extent of accumulation of ApoER2-Dab1 pathway components in amygdala correlated with the presence or extent of the neurobehavioral endpoints in the NACC Uniform Data Set: behavior, comportment, and personality (Form B4, Item 9); personality change (Form B9, Item 9g); behavioral symptoms (Form B9, Item 8); and depression (Form B6, Item 16; GDS-15). Corresponding analyses were completed in ErC to examine the potential specificity for any correlations observed between amygdala pathology and neuropsychiatric endpoints. Deficits in comportment showed the strongest and most robust associations with ApoER2-Dab1 pathway components (**Fig 7**). In amygdala, the extent of accumulation of all seven ApoER2-Dab1 pathway markers that were significantly elevated in AD—ApoER2, Dab1, pP85α_Tyr607_, pLIMK1_Thr508_, pTau_Ser202/Thr205_, pPSD95_Thr19_, and ApoJ— correlated with deficits in comportment. In ErC, accumulations of four pathway components (pP85α_Tyr607_, pTau_Ser202/Thr205_, pPSD95_Thr19_, ApoJ,) were similarly correlated with comportment. Associations of ApoER2-Dab1 pathway components with behavioral symptoms, personality changes, and depression were present but less robust. Pathway components tended to be positively associated with behavioral symptoms and personality changes and inversely associated with depression, with stronger associations observed in the amygdala than in ErC. The observed inverse correlations between neuronal pathway components and depression score were unexpected. Since AD severity is associated with apathy,[40] lack of interest or concern could potentially mitigate affective responses to cognitive and functional deficits in AD.

## 4 Discussion

Emotional dysregulation and mood disorders are common manifestations of AD that have major negative consequences for patients and caregivers. The amygdala—which plays a central role in the regulation of emotions, behavior, and mood—is severely affected by both neuritic plaques and NFTs in advanced AD [12, 13, 15, 17, 41–43] and is thought to degenerate early in a subset of AD patients. However, specific mechanisms underlying degeneration of the amygdala have not yet been identified, and treatments for these neuropsychiatric manifestations of AD are currently limited. We previously showed that multiple ApoER2-Dab1 pathway components accumulate together in five regions that degenerate in the earliest stages of AD,[25] and two regions that degenerate later in the disease process,[27] and proposed that ApoER2-Dab1 disruption could potentially be a universal mechanism underlying AD-related degeneration in humans. In the present study in amygdala, we observed that: (1) ApoER2 is highly expressed by a subset or neurons in cognitively normal controls; (2) ApoER2 accumulates together with five of its neuronal signaling partners (Dab1, pP85α_Tyr607_, pLIMK1_Thr508_, pTau _Ser202/Thr205_ and pPSD95_Thr19_) and one of its extracellular ligands (ApoJ) within abnormal neurons and extracellular plaques, respectively, in MCI and AD cases; and (3) these accumulations of ApoER2-Dab1 pathway components correlated with histological progression, cognitive deficits, and neuropsychiatric endpoints. Findings add to growing evidence implicating ApoER2-Dab1 pathway disruption in AD-related neurodegeneration, and suggest that compromised function of the ApoER2-Dab1 pathway in amygdala could contribute to cognitive and neuropsychiatric manifestations of AD.

### 4.1 Could ApoER2-Dab1 pathway dysfunction be a universal mechanism underlying neuro-degeneration in AD?

An extensive body of preclinical studies demonstrated that ApoER2 plays essential roles in memory, cognition, and neuronal integrity (reviewed in [25, 27]) and that signaling through the ApoER2-Dab1 pathway regulates both Tau phosphorylation [21–25, 44] and Aβ production.[45–47] Using rapidly autopsied specimens from seven human brain regions that are vulnerable to AD pathology—entorhinal cortex, locus coeruleus, raphe nucleus, prosubiculum-CA1 border region, temporal neocortex, hippocampus, molecular layer of dentate gyrus [25, 27]—we previously reported that: (1) ApoER2 and *LRP8* (gene encoding ApoER2 protein) are strongly expressed in the same regions, layers, and neuron populations that develop pTau pathology in the earliest stages of AD; (2) the same neurons that accumulate pTau in early AD strongly express ApoER2; and (3) ApoER2 accumulates together with five of its neuronal signaling partners (Dab1, pP85α_Tyr607_, pLIMK1_Thr508_, pTau_Ser202/Thr205_ and pPSD95_Thr19_) and three extracellular ApoER2 ligands (ApoE, ApoJ, Reelin) in abnormal neurons and neuritic plaques, respectively, in MCI and AD cases. These collective findings provided the basis for an alternative mechanistic paradigm and unifying model implicating disruption of the ApoER2-Dab1 pathway in AD pathogenesis. This model is attractive because it integrates hallmark pathologies such as pTau, Aβ, ApoJ and ApoE, with emerging AD pathologies such as Reelin, Dab1, and ApoER2 into a coherent model. Important aspects of this model are bolstered by evidence that: (1) a gain-of-function variant in the *RELN* gene protects from familial, autosomal dominant AD in humans,[48, 49] (2) the *DAB1* gene locus is associated with AD risk in *APOE* ε4 homozygotes,[50] and (3) a single-cell transcriptomics report suggesting that vulnerable neuron subpopulations have higher expression of *RELN*, *DAB1* and *LRP8* than less vulnerable neurons.[51] Altogether, these findings support the concept that disruption or compromised function of the ApoER2-Dab1 pathway could contribute to neurodegeneration in AD. Importantly, however, it is not yet known whether ApoER2-Dab1 disruption contributes to the degeneration of the amygdala or the neuropsychiatric manifestations of AD.

### 4.2 Does ApoER2-Dab1 pathway disruption underlie amygdala degeneration in AD?

The amygdala has been known to undergo severe neurodegeneration in AD since 1938.[12, 15–17, 41–43] In the early 1990s, Kromer [15] and Hyman [12] showed that hallmark Aβ-containing plaques and NFTs preferentially accumulate within subregions connecting the amygdala to hippocampus and ErC, and proposed that ensuing disruptions could underlie cognitive and neuropsychiatric aspects of AD. In the present study, our finding that seven ApoER2-Dab1 pathway components accumulate in amygdala and correlate with comportment, personality, and depression, suggest that ApoER2-Dab1 pathway disruption may contribute to the complex neuropsychiatric manifestations of AD. As previously observed in hippocampus and the molecular layer of dentate gyrus,[27] and in the ErC, prosubiculum-CA1 border region, middle temporal gyrus, locus coeruleus and raphe nucleus,[25] multiplex-IHC in amygdala revealed that multiple neuronal ApoER2-Dab1 pathway markers (ApoER2, Dab1, pP85α_Tyr607_, pLIMK1_Thr508_, pTau_Ser202/Thr205_, PSD95_Thr19_) accumulated in the soma and MAP2-labeled dystrophic dendrites of abnormal neurons. As previously described in hippocampus,[25, 27] Dab1 was unique among these ApoER2 pathway components because it also accumulated within a subset of NFL-labeled dystrophic axons, which were particularly prominent in the molecular layers of the dentate gyrus and cornu ammonis. Since Dab1 is a cytoplasmic adaptor protein for both ApoER2 and AβPP [52, 53] and ApoER2 signaling has been reported to modulate AβPP cleavage in model systems,[45–47] the accumulation of Dab1 within dystrophic axons surrounding ApoE-Aβ plaques suggests that axonal ApoER2-Dab1 disruption may regulate Aβ synthesis in humans. When considered together with evidence from model systems that ApoER2-Dab1 signaling suppresses Tau phosphorylation,[54–57] these findings lend support to our unifying model wherein ApoER2 disruption could be a shared molecular origin linking ApoE, ApoJ, Reelin and Dab1 to the Aβ plaques and pTau tangles that define AD in humans (reviewed in [25, 27, 58]).

### 4.3 Conflicting role of Reelin in amygdala and hippocampus?

Reelin is reported to play an important role in development of amygdala in mice;[59] however, it is not yet known whether Reelin plays a role in maintaining the integrity of the human amygdala in adulthood, or in the pathogenesis of AD. Our previous work showed prominent neuritic plaque-associated deposition of Reelin in the molecular layer of the dentate gyrus and cornu ammonis in most (but not all) AD cases.[27] Unexpectedly, however, despite using the same reagents and methods on corresponding amygdala specimens, we did not find evidence for Reelin accumulation in the amygdala in any of these same AD cases. This difference suggests that different molecular mechanisms may compromise ApoER2-Dab1 pathway function in different brain regions. Since Reelin signaling through ApoER2 induces proteasomal degradation of Dab1 (reviewed in [25, 27]), the observed combination of extracellular Reelin deposition and neuritic Dab1 accumulation in hippocampus implies that disruption of Reelin binding to ApoER2 in hippocampus may play a role in AD pathogenesis. By contrast, the presence of neuritic Dab1 accumulation in amygdala despite the lack of extracellular Reelin accumulation in the present study implies that Reelin depletion may play a more prominent role in AD pathogenesis in amygdala. However, future studies are needed to clarify the specific molecular mechanisms underlying ApoER2-Dab1 pathway dysfunction in amygdala.

### 4.4 Strengths & limitations

The use of rapidly-autopsied specimens that underwent uniform, time-limited fixation in conjunction with antemortem cognitive and neuropsychiatric data, and the pathological and cytoarchitectural context provided by MP-IHC are important strengths of the present study.

Postmortem specimens spanning the clinicopathological spectrum of AD were used to approximate AD progression. This cross-sectional study design cannot establish a sequence of disease progression. Preclinical studies with rodent models or human iPSC-derived neurons are needed to elucidate the roles of individual ApoER2-Dab1 pathway components in neurodegeneration and AD. The moderate sample size is an important limitation; larger studies using specimens collected from additional cohorts will provide additional insights and can determine if results are influenced by sex, genetics, or other variables. The amygdala is a highly complex and heterogenous structure comprising approximately 13 nuclei;[60] thus, future studies parsing these individual nuclei may provide insights into region-specific vulnerability to ApoER2-Dab1 pathway disruption. MP-IHC was performed on a subset of representative cases with 20x magnification. Future studies using higher resolution imaging are needed to determine whether ApoER2-Dab1 pathway markers accumulate and co-localize within neuronal lysosomes, autophagosomes, proteasomes, and other organelles.

### 4.5 Summary & Conclusion

We found that (1) ApoER2 is highly expressed by a subset of amygdala neurons; (2) multiple ApoER2-Dab1 pathway components accumulate within abnormal neurons and neuritic plaques in the amygdala in MCI and AD; and (3) ApoER2-Dab1 pathway markers correlate with hallmark plaques and tangles, cognition, and neuropsychiatric manifestations of AD. These findings strengthen and extend the concept that ApoER2-Dab1 pathway disruption may be a universal mechanism underlying AD-related degeneration, and suggest that ApoER2-Dab1 disruption in amygdala may contribute to the neuropsychiatric manifestations of AD.

## Abbreviations

AD: Alzheimer’s disease
ApoE: Apolipoprotein E
ApoER2: ApoE receptor 2
ApoJ: Apolipoprotein J
Dab1: Disabled homolog-1
DAPI: nuclear marker
GFAP: Glial fibrillary acidic protein
HIER: Heat induced epitope retrieval
IBA1: ionized calcium-binding adapter molecule 1
LIMK1: LIM domain kinase-1
*LRP8*: Low-density lipoprotein receptor-related protein 8 (gene encoding ApoER2)
MAP2: Microtubule associated protein 2
NEUN: neuronal nuclear/soma antigen
NFL: Neurofilament light chain
P85α: PI3K regulatory subunit P85alpha
PMI: Postmortem interval
PSD95: Postsynaptic density protein 95
pTau: phosphorylated Tau_Ser202/Thr205_
*RELN*: gene encoding Reelin protein
sAD: sporadic AD
SYNP: Synaptophysin

## Data Availability

All data produced in the present study are available upon reasonable request to the authors

## Acknowledgements

We are grateful to the research volunteers and the BBDP research team for the provision of human biological materials, and Jahandar Jahanipour for his contribution to post-acquisition image processing.

## 8 Additional file

- File name: supplement.docx
- Title: Supplement
- Description: Contains supplementary tables, extended figures, and supplementary text on materials and methods.

