## supplement for "Evidence for ApoE receptor 2-Disabled homolog-1 pathway disruption in the amygdala in sporadic Alzheimer’s disease"

**I. Supplementary Tables**

| **Table** | **Title** |
| --- | --- |
| S1 | Thirty-two cases spanning the clinicopathological spectrum of Alzheimer's disease progression. |
| S2 | Summary characteristics (n=32). |
| S3 | Description of NACC Uniform Data Set variables. |
| S4 | Key resources. |
| S5 | False discovery rate adjusted p-values for manuscript figures. |

**Table S1. Thirty-two cases spanning the clinicopathological spectrum of Alzheimer's disease progression.**

| **ID** | **Sex** | **Age** | **PMI** | **Braak** | **Thal** | **Amyloid** | **Neuritic** | **MMSE** | **ApoE** |
| --- | --- | --- | --- | --- | --- | --- | --- | --- | --- |
|  |  | **(years) ᵃ** | **(hours)** | **stage** | **phase** | **plaques** | **plaques** | **(0-30)** | **status** |
|  |  |  |  | **(0-VI)** | **(0-5)** | **(0-15) ᵇ** |  |  |  |
| **Alzheimer's Disease** | | | | | | | | | |
| 1 | Male | 70-74 | 4.8 | VI | 5 | 14 | Frequent | 7 | 3/4 |
| 2 | Female | 70-74 | 4.9 | VI | 4 | 15 | Frequent | 17 | 3/4 |
| 3 | Female | 75-79 | 3.1 | V |  | 15 | Frequent | 6 | 3/3 |
| 4 | Male | 75-79 | 3.6 | VI | 5 | 15 | Frequent | 10 | 3/4 |
| 5 | Male | 80-84 | 4.0 | V | 5 | 14 | Frequent | 14 | 3/4 |
| 6 | Female | 80-84 | 4.0 | VI | 5 | 15 | Frequent | 19 | 3/3 |
| 7 | Female | 80-84 | 3.3 | VI | 5 | 15 | Frequent | 24 | 3/3 |
| 8 | Male | 85-89 | 2.2 | V |  | 12 | Frequent | 21 | 2/3 |
| 9 | Female | 85-89 | 3.4 | VI | 5 | 14 | Frequent | 2 | 3/3 |
| 10 | Female | ≥90 | 2.2 | V | 3 | 15 | Frequent | 13 | 3/3 |
| **Mild Cognitive Impairment** | | | | | | | | | |
| 11 | Female | 75-79 | 3.2 | IV | 2 | 4 | None | 24 | 2/3 |
| 12 | Female | 80-84 | 3.0 | IV |  | 12 | Frequent | 29 | 2/3 |
| 13 | Female | 80-84 | 3.2 | IV | 4 | 11 | Frequent | 29 | 2/3 |
| 14 | Male | 85-89 | 2.2 | III | 5 | 8 | Moderate | 28 | 3/3 |
| 15 | Male | 85-89 | 4.2 | IV | 0 | 0 | None | 23 | 3/3 |
| 16 | Female | 85-89 | 3.6 | IV | 3 | 10 | Frequent | 28 | 2/3 |
| 17 | Female | ≥90 | 3.2 | IV | 2 | 5 | Frequent | 22 | 3/3 |
| 18 | Female | ≥90 | 3.2 | IV |  | 14 | Frequent | 26 | 2/3 |
| **Age-matched Controls** | | | | | | | | | |
| 19 | Male | 70-74 | 4.6 | I | 0 | 0 | None | 29 | 3/3 |
| 20 | Male | 70-74 | 3.5 | III | 0 | 0 | None | 27 | 3/3 |
| 21 | Male | 75-79 | 2.3 | I | 2 | 6 | Sparse | 29 | 3/4 |
| 22 | Female | 80-84 | 2.1 | I | 1 | 1 | Sparse | 29 | 2/3 |
| 23 | Male | 80-84 | 2.0 | I | 2 | 0 | None |  | 3/3 |
| 24 | Male | 85-89 | 3.0 | I | 2 | 4 | Sparse |  | 3/3 |
| 25 | Female | 85-89 | 3.1 | III | 0 | 0 | None | 28 | 3/4 |
| 26 | Male | ≥90 | 3.4 | I | 1 | 0 | Sparse | 27 | 3/3 |
| 27 | Female | ≥90 | 3.0 | III | 3 | 6 | Sparse | 27 | 3/3 |
| **Middle-age Controls** | | | | | | | | | |
| 28 | Male | 35-39 | 3.0 | 0 | 0 | 0 | None |  | 3/3 |
| 29 | Male | 45-49 | 4.5 | 0 | 0 | 0 | None |  | 3/3 |
| 30 | Female | 50-54 | 4.7 | I | 0 | 0 | None |  | 3/3 |
| 31 | Female | 55-59 | 3.1 | I | 1 | 1 | None |  | 3/3 |
| 32 | Male | 60-64 | 2.3 | I | 1 | 0 | Sparse |  | 3/3 |
| ᵃ Ages of individual cases are presented in 5-year intervals to ensure patient confidentiality. In addition, BBDP classified all cases that were at least 90 years of age as "≥90". | | | | | | | | | |
| ᵇ Amyloid plaque density or neurofibrillary tangle density score in the following regions: frontal, temporal, parietal, hippocampus, and entorhinal cortex. | | | | | | | | | |

**Table S2. Summary characteristics (n=32) ᵃ**

|  | **AD (n=10)** | **MCI (n=8)** | **Control (n=9)** | **Young Control (n=5)** |
| --- | --- | --- | --- | --- |
| **Demographic and clinical characteristics** |  |  |  |  |
| Age, y, median (range) ᵇ | 82 (73-90) | 85 (76-90) | 84 (71-90) | 52 (38-61) |
| Education, y, median (range) | 14 (12-16) | 13 (12-16) | 14 (10-18) | 16 (14-19) |
| Female | 6 | 6 | 3 | 2 |
| Post-mortem interval, hours, median (range) | 3.5 (2.2-4.9) | 3.2 (2.2-4.2) | 3.0 (2.0-4.6) | 3.1 (2.3-4.7) |
| **Neuropathology and genetics** |  |  |  |  |
| Thal phase (0-5), mean (range) | 5 (3-5) | 2 (0-5) | 1 (0-3) | 0 (0-1) |
| Braak stage (0-6), median (range) | 6 (5-6) | 4 (3-4) | 1 (1-3) | 1 (0-1) |
| Neurofibrillary tangle (0-15), median (range) ᶜ | 15 (10-15) | 7 (5-8) | 2 (0-4) | 0 (0-0) |
| Entorhinal cortex (0-3), median (range) | 3 (3-3) | 3 (2-3) | 1 (0-2) | 0 (0-0) |
| Hippocampus (0-3), median (range) | 3 (2-3) | 3 (2-3) | 0 (0-2) | 0 (0-0) |
| Temporal cortex (0-3), median (range) | 3 (2-3) | 1 (0-2) | 0 (0-1) | 0 (0-0) |
| Neuritic plaque density (0-3), median (range) | 3 (3-3) | 3 (0-3) | 1 (0-1) | 0 (0-1) |
| Amyloid plaques (0-15), median (range) ᶜ | 15 (12-15) | 9 (0-14) | 0 (0-6) | 0 (0-1) |
| Entorhinal cortex (0-3), median (range) | 3 (2-3) | 2 (0-3) | 0 (0-2) | 0 (0-0) |
| Hippocampus (0-3), median (range) | 3 (1-3) | 0 (0-2) | 0 (0-1) | 0 (0-0) |
| Temporal cortex (0-3), median (range) | 3 (3-3) | 2 (0-3) | 0 (0-2) | 0 (0-1) |
| **Cognitive endpoints** |  |  |  |  |
| Cognitive dysfunction, y, median (range) | 8 (1-15) |  |  |  |
| MMSE (0-30), median (range) | 14 (2-24) | 27 (22-29) | 28 (27-29) |  |
| Clinical Dementia Rating sum of boxes (0-18), median (range) | 13 (0-18) | 0 (0-2) | 0 (0-0) |  |
| Clinical Dementia Rating global score (0-3), median (range) | 2 (0-3) | 0 (0-0) | 0 (0-0) |  |
| FAST Score (1-7), median (range) | 4 (1-6) | 2 (1-3) | 1 (1-2) |  |
| Figure Recall Score (0-3), median (range) | 1 (0-2) | 2 (1-2) | 3 (3-3) |  |
| AVLT Total Learning (0-75), median (range) ᶜ | 21 (12-46) | 48 (21-48) | 43 (25-52) |  |
| AVLT STM A6 (0-15), median (range) | 4 (0-8) | 9 (4-10) | 10 (8-11) |  |
| WMSR Digit Span Forward Score (0-12), median (range) | 8 (6-9) | 8 (6-12) | 10 (7-11) |  |
| ApoE, n |  |  |  |  |
| 3/3 | 5 | 3 | 6 | 5 |
| 2/3 | 1 | 5 | 1 | 0 |
| 3/4 | 4 | 0 | 2 | 0 |
| NIA-Reagan, Likelihood of Alzheimer's disease, n ᵈ |  |  |  |  |
| Not AD | 0 | 0 | 1 | 0 |
| Intermediate | 0 | 1 | 0 | 0 |
| High | 10 | 0 | 0 | 0 |
| Criteria Not Met | 0 | 7 | 8 | 5 |
| Dementia not otherwise specified, n | 0 | 0 | 0 | 0 |
| Hippocampal sclerosis, n | 0 | 0 | 0 | 0 |
| Vascular dementia, n ᵉ | 0 | 0 | 0 | 0 |
| ᵃ Some markers have fewer than 30 cases (no less than 21). ᵇ BBDP classified all cases >90 years of age as "90 years of age" to ensure confidentiality. ᶜ Total scores for plaques and tangles include the entorhinal, hippocampus, temporal, parietal, and frontal areas. Each was scored according to the CERAD templates [1] using Campbell-Switzer silver stain, Gallyas silver stain and Thioflavin S stains. ᵈ Modified NIA-Reagan diagnosis of Alzheimer's disease based on consensus recommendations for postmortem diagnosis of Alzheimer's disease [2] including neurofibrillary tangles (Braak) and neuritic plaques (CERAD). ᵉ Defined by NINDS-AIREN criteria.[3] | | | | |

| **Table S3. Description of NACC Uniform Data Set variables** | | | | |
| --- | --- | --- | --- | --- |
| **Name in figures** | **Form** | **Question No.** | **Short descriptor** | **Question/Description** |
| Comportment | B4 CDR Plus NACC FTLD | 9 | Behavior, comportment, and personality | This domain is intended to assess changes in personality, aberrant behaviors, and changes in interpersonal relationships. The kinds of specific issues that might fall under these rubrics include: loss of insight, disinhibition, apathy, social withdrawal and disengagement, emotional lability, easy distractibility, reduced empathy for the feelings of others, impulsivity, and changes in eating habits and table manners. A key element of each of these is the degree to which these behaviors impact interpersonal relationships. |
| Total severity | B5 Neuropsychiatric Inventory Questionnaire (NPI-Q) | N/A | Sum of all severity questions in NPI-Q | A brief assessment of neuropsychiatric symptomatology in routine clinical practice settings. The total score is for 12 questions. |
| Apathy (NPIQ) | B5 Neuropsychiatric Inventory Questionnaire (NPI-Q) | 8 | Apathy or indifference severity | Does the patient seem less interested in his/her usual activities or in the activities and plans of others? |
| Depression | B6 Geriatric Depression Scale (GDS) | N/A | Total GDS Score | Basic screening measure for depression in older adults. The total score is for 15 questions. |
| Behavioral symptoms | B9 Clinician Judgment of Symptoms | 8 | Based on clinician’s judgment, is the subject currently experiencing any kind of behavioral symptoms? | Decline or changes in behavior refers to meaningful change or decline from the subject’s usual or customary behavior reported or observed at the current visit. |
| Apathy (CJS) | B9 Clinician Judgment of Symptoms | 9a | Subject currently manifests meaningful change in behavior — Apathy, withdrawal | Has the subject lost interest in or displayed a reduced ability to initiate usual activities and social interaction, such as conversing with family and/or friends? |
| Personality change | B9 Clinician Judgment of Symptoms | 9g | Subject currently manifests meaningful change in behavior — Personality change | Does the subject exhibit bizarre behavior or behavior uncharacteristic of the subject, such as unusual collecting, suspiciousness (without delusions), unusual dress, or dietary changes? Does the subject fail to take others’ feelings into account? |
| Adapted from the NACC Uniform Data Set (UDS) Researcher's Data Dictionary as well as the Coding Guidebook for Initial Visit Packet (Version 3.0, March 2015, for both documents) | | | | |

**Table S4. Key Resources**

| ***Reagent type or resource*** | ***Target*** | ***Designation*** | ***Type*** | ***Source*** | ***Catalog #*** | ***Previous uses and validation*** | ***IHC conditions*** |
| --- | --- | --- | --- | --- | --- | --- | --- |
| **Primary antibodies for IHC** | |  |  |  |  |  |  |
| antibody | ApoE receptor 2 | ApoER2 | Rabbit IgG | Millipore-Sigma | SAB2103110 | IHC human brain FFPE, positive and negative control IHC, WB in OE vs Control lysates, multi-epitope labeling | pH6 at 70C for 40 min; 1:80-1:150 |
| antibody | Disabled homolog 1 | Dab1 | Rabbit IgG | Invitrogen | PA5-86617 | IHC human brain FFPE, positive and negative control IHC, WB in OE vs Control lysates, multi-epitope labeling | pH6 at 70C for 40 min; 1:50 |
| antibody | Tyr607-phosphorylated P85α | pP85α_Tyr607_ | Rabbit IgG | Invitrogen | PA5-104853 | IHC human brain FFPE, positive and negative control IHC, WB, WB with phospho-blocking peptide | pH6 at 70C for 40 min; 1:100-1:150 |
| antibody | Thr508-phosphorylated LIM kinase-1 | pLIMK1_Thr508_ | Rabbit IgG | Invitrogen | PA5-104925 | IHC human brain FFPE, positive and negative control IHC, WB, WB with phospho-blocking peptide | pH6 at 70C for 20 min; 1:100 |
| antibody | Ser202/Thr205-phosphorylated Tau | pTau | Mouse IgG1 [AT8] | Invitrogen | MN1020 | IHC human brain FFPE, WB | pH6 at 70C for 40 min; 1:100 |
| antibody | Thr19-phosphorylated PSD95 | pPSD95_Thr19_ | Rabbit IgG | Millipore-Sigma | ABN998 | IHC human brain FFPE, positive and negative control IHC, WB in OE vs Control lysates, multi-epitope labeling | pH6 at 70C for 40 min; 1:50 |
| antibody | Apolipoprotein E | ApoE | Mouse IgG1 [WUE4] | Novus Biologicals | NB110-60531 | IHC human brain FFPE, positive and negative control IHC, WB, multi-epitope labeling | pH6 at 70C for 40 min; 1:60-1:100 |
| antibody | Apolipoprotein E | ApoE | Chicken IgG | Biosynth | NEP4809 | IHC human brain FFPE, positive and negative control IHC, WB, multi-epitope labeling | 88% formic acid for 10 min, 1:50 |
| antibody | Apolipoprotein J | ApoJ | Rabbit IgG | Invitrogen | PA5-24426 | IHC human brain FFPE, positive and negative control IHC, WB | pH6 at 70C for 40 min; 1:100-1:150 |
| antibody | Reelin | Reelin | Mouse IgG2a [E-5] | Santa Cruz | SC-25346 | IHC human brain FFPE, positive and negative control IHC, WB | pH6 at 70C for 25 min; 1:50 |
| antibody | Amyloid Beta Protein | Aβ | Mouse IgG2b [MOAB-2] | Novus Biologicals | NBP2-13075 | IHC human brain FFPE, positive and negative control IHC, WB, multi-epitope labeling | 88% formic acid for 10 min, 1:100 |
| **Cytoarchitectural antibodies for multiplex-IHC** | |  |  |  |  |  |  |
| antibody | Neuronal marker NeuN | NEUN | Guinea Pig IgG | Millipore Sigma | ABN90P | IHC human brain FFPE, positive and negative control IHC, WB | pH9 at 70C for 40 min; 1:100 |
| antibody | Microtubule associated protein 2 | MAP2 | Mouse IgG3 [885232] | R&D Systems | MAB8304 | IHC human brain FFPE, positive and negative control IHC | pH9 at 70C for 40 min; 1:100 |
| antibody | Glial fibrillary acidic protein | GFAP | Rat IgG2a [2.2B10] | Invitrogen | 13-0300 | IHC human brain FFPE, positive and negative control IHC | pH9 at 70C for 40 min; 1:100 |
| antibody | Ionized calcium-binding adaptor molecule 1 | IBA1 | Guinea Pig IgG | Synaptic Systems | 234005 | IHC human brain FFPE | pH9 at 70C for 20 min; 1:100 |
| antibody | Neurofilament light chain | NFL | Mouse IgG1 [NFL2] | Biolegend | 846002 | IHC human brain, positive and negative control IHC, WB, multi-epitope labeling | pH9 at 70C for 40 min; 1:100 |
| antibody | Neurofilament light chain | NFL | Mouse IgG1 [NFL3] | Biolegend | 845902 | IHC human brain, positive and negative control IHC, WB, multi-epitope labeling | pH9 at 70C for 40 min; 1:100 |
| antibody | Synaptophysin | SYNP | Mouse IgM [SP15] | Millipore Sigma | MAB329 | IHC human brain FFPE, positive and negative control IHC, WB | pH9 at 70C for 40 min; 1:100 |
| **Secondary Antibodies for IHC** | |  |  |  |  |  |  |
| antibody | Goat anti-Rat IgG |  |  | Jackson | 112-035-167 |  | 1.6 ug/mL |
| antibody | Goat anti-Mouse IgG1 |  |  | Jackson | 115-035-205 |  | 1.6 ug/mL |
| antibody | Goat anti-Mouse IgG2a |  |  | Jackson | 115-035-206 |  | 1.6 ug/mL |
| antibody | Goat anti-Mouse IgG2b |  |  | Jackson | 115-035-207 |  | 1.6 ug/mL |
| antibody | Goat anti-Mouse IgG3 |  |  | Jackson | 115-035-209 |  | 1.6 ug/mL |
| antibody | Goat anti-Mouse IgM |  |  | Jackson | 115-035-075 |  | 1.6 ug/mL |
| antibody | Donkey anti-Rabbit IgG |  |  | Jackson | 711-035-152 |  | 1.6 ug/mL |
| Abbreviations: IHC, immunohistochemistry; HIER, heat induced epitope retrieval; FFPE, formalin-fixed paraffin embedded; WB, western blot; OE, overexpression. | | | | | | | |

| **Table S5. False discovery rate adjusted p-values for manuscript figures ᵃ** | | | | | | | | | | | | | | | |
| --- | --- | --- | --- | --- | --- | --- | --- | --- | --- | --- | --- | --- | --- | --- | --- |
|  |  | **by Group** | |  | **vs Braak stage** | |  | **vs Amyloid plaques** | |  | **vs MMSE** | |  | **vs CAA total** | |
|  |  | **p-value ᵇ** | **Sharpened** |  | **p-value ᶜ** | **Sharpened** |  | **p-value ᶜ** | **Sharpened** |  | **p-value ᶜ** | **Sharpened** |  | **p-value ᶜ** | **Sharpened** |
|  |  |  | **q-value** |  |  | **q-value** |  |  | **q-value** |  |  | **q-value** |  |  | **q-value** |
| **Figure 1.B** | | | | | | | | |  |  |  |  |  |  |  |
| ApoER2 | Amygdala | 0.009 | 0.020 |  | 0.008 | 0.020 |  | 0.003 | 0.009 |  | 0.020 | 0.035 |  | 0.096 | 0.081 |
| **Figure 2.B** | | | | | | | | |  |  |  |  |  |  |  |
| Dab1 | Amygdala | 0.006 | 0.017 |  | 0.006 | 0.017 |  | 0.004 | 0.013 |  | 0.052 | 0.060 |  | 0.008 | 0.020 |
| pP85α | Amygdala | <0.001 | 0.001 |  | <0.001 | 0.001 |  | <0.001 | 0.001 |  | <0.001 | 0.001 |  | <0.001 | 0.002 |
| pLIMK1 | Amygdala | 0.001 | 0.005 |  | 0.003 | 0.010 |  | <0.001 | 0.004 |  | 0.001 | 0.005 |  | <0.001 | 0.002 |
| **Figure 3.B** | | | | | | | | |  |  |  |  |  |  |  |
| pTau | Amygdala | <0.001 | 0.001 |  | <0.001 | 0.001 |  | <0.001 | 0.001 |  | <0.001 | 0.001 |  | <0.001 | 0.002 |
| pPSD95 | Amygdala | <0.001 | 0.001 |  | <0.001 | 0.001 |  | <0.001 | 0.001 |  | <0.001 | 0.001 |  | <0.001 | 0.001 |
| **Figure 5.B** | | | | | | | | |  |  |  |  |  |  |  |
| ApoJ | Amygdala | <0.001 | 0.002 |  | <0.001 | 0.001 |  | <0.001 | 0.002 |  | <0.001 | 0.002 |  | 0.002 | 0.007 |
| ApoE | Amygdala | 0.429 | 0.209 |  | 0.210 | 0.139 |  | 0.128 | 0.102 |  | 0.774 | 0.330 |  | 0.076 | 0.074 |
| Reelin | Amygdala | 0.786 | 0.330 |  | 0.530 | 0.255 |  | 0.280 | 0.168 |  | 0.709 | 0.303 |  | 0.867 | 0.369 |
|  |  |  |  |  | **vs Comportment** | |  | **vs Behavioral symptoms** | |  | **vs Personality change** | |  | **vs GDS-15** | |
|  |  |  |  |  | **p-value ᶜ** | **Sharpened** |  | **p-value ᵈ** | **Sharpened** |  | **p-value ᵈ** | **Sharpened** |  | **p-value ᶜ** | **Sharpened** |
|  |  |  |  |  |  | **q-value** |  |  | **q-value** |  |  | **q-value** |  |  | **q-value** |
| **Figure 7.A** | | | | | | | | |  |  |  |  |  |  |  |
| ApoER2 | Amygdala |  |  |  | 0.033 | 0.048 |  | 0.158 | 0.113 |  | 0.083 | 0.077 |  | 0.082 | 0.077 |
|  | ErC |  |  |  | 0.142 | 0.106 |  | 0.037 | 0.049 |  | 0.386 | 0.201 |  | 0.292 | 0.169 |
| Dab1 | Amygdala |  |  |  | 0.021 | 0.037 |  | 0.024 | 0.038 |  | 0.248 | 0.156 |  | 0.065 | 0.068 |
|  | ErC |  |  |  | 0.534 | 0.255 |  | 0.365 | 0.190 |  | 0.665 | 0.296 |  | 0.557 | 0.261 |
| pP85α | Amygdala |  |  |  | 0.001 | 0.005 |  | 0.017 | 0.032 |  | 0.194 | 0.132 |  | 0.011 | 0.024 |
|  | ErC |  |  |  | <0.001 | 0.002 |  | 0.024 | 0.038 |  | 0.043 | 0.056 |  | 0.084 | 0.077 |
| pLIMK1 | Amygdala |  |  |  | 0.006 | 0.017 |  | 0.013 | 0.026 |  | 0.312 | 0.173 |  | 0.029 | 0.044 |
|  | ErC |  |  |  | 0.094 | 0.081 |  | 0.024 | 0.038 |  | 0.665 | 0.296 |  | 0.468 | 0.229 |
| pTau | Amygdala |  |  |  | 0.001 | 0.005 |  | 0.051 | 0.060 |  | 0.030 | 0.045 |  | 0.036 | 0.049 |
|  | ErC |  |  |  | 0.005 | 0.015 |  | 0.069 | 0.070 |  | 0.112 | 0.093 |  | 0.045 | 0.057 |
| pPSD95 | Amygdala |  |  |  | 0.002 | 0.006 |  | 0.037 | 0.049 |  | 0.030 | 0.045 |  | 0.025 | 0.039 |
|  | ErC |  |  |  | 0.007 | 0.017 |  | 0.051 | 0.060 |  | 0.112 | 0.093 |  | 0.025 | 0.039 |
| **Figure 7.B** | | | | | | | | |  |  |  |  |  |  |  |
| ApoJ | Amygdala |  |  |  | 0.016 | 0.030 |  | 0.312 | 0.173 |  | 0.398 | 0.203 |  | 0.060 | 0.064 |
|  | ErC |  |  |  | 0.009 | 0.020 |  | 0.061 | 0.064 |  | 0.091 | 0.081 |  | 0.345 | 0.183 |
| ApoE | Amygdala |  |  |  | 0.784 | 0.330 |  | 0.628 | 0.284 |  | 0.586 | 0.270 |  | 0.736 | 0.314 |
|  | ErC |  |  |  | 0.363 | 0.190 |  | 0.628 | 0.284 |  | 0.697 | 0.300 |  | 0.259 | 0.156 |
| Reelin | Amygdala |  |  |  | 0.338 | 0.183 |  | 0.053 | 0.060 |  | 0.346 | 0.183 |  | 0.649 | 0.293 |
|  | ErC |  |  |  | 0.877 | 0.370 |  | 0.053 | 0.060 |  | 0.814 | 0.342 |  | 0.601 | 0.276 |
| ᵃ Based on the two-stage linear step-up procedure described by Benjamini, et al. and Anderson.[4, 5] | | | | | | | | | | | | | | | |
| ᵇ Determined using the Kruskal-Wallis equality-of-populations rank test. | | | | | | | | | | | | | | | |
| ᶜ Determined using the Spearman's rank correlations test. | | | | | | | | | | | | | | | |
| ᵈ Determined using the Wilcoxon rank-sum test. | | | | | | | | | | | | | | | |
| Abbreviations: MMSE, Mini-Mental State Examination; CAA, cerebral amyloid angiopathy; GDS, Geriatric Depression Scale. | | | | | | | | | | | | | | | |

**II. Extended Figures**

| **Ext Fig** | **Title** |
| --- | --- |
| 7 | Correlations between ApoER2-Dab1 pathway components with depression, apathy and total NPI-Q |

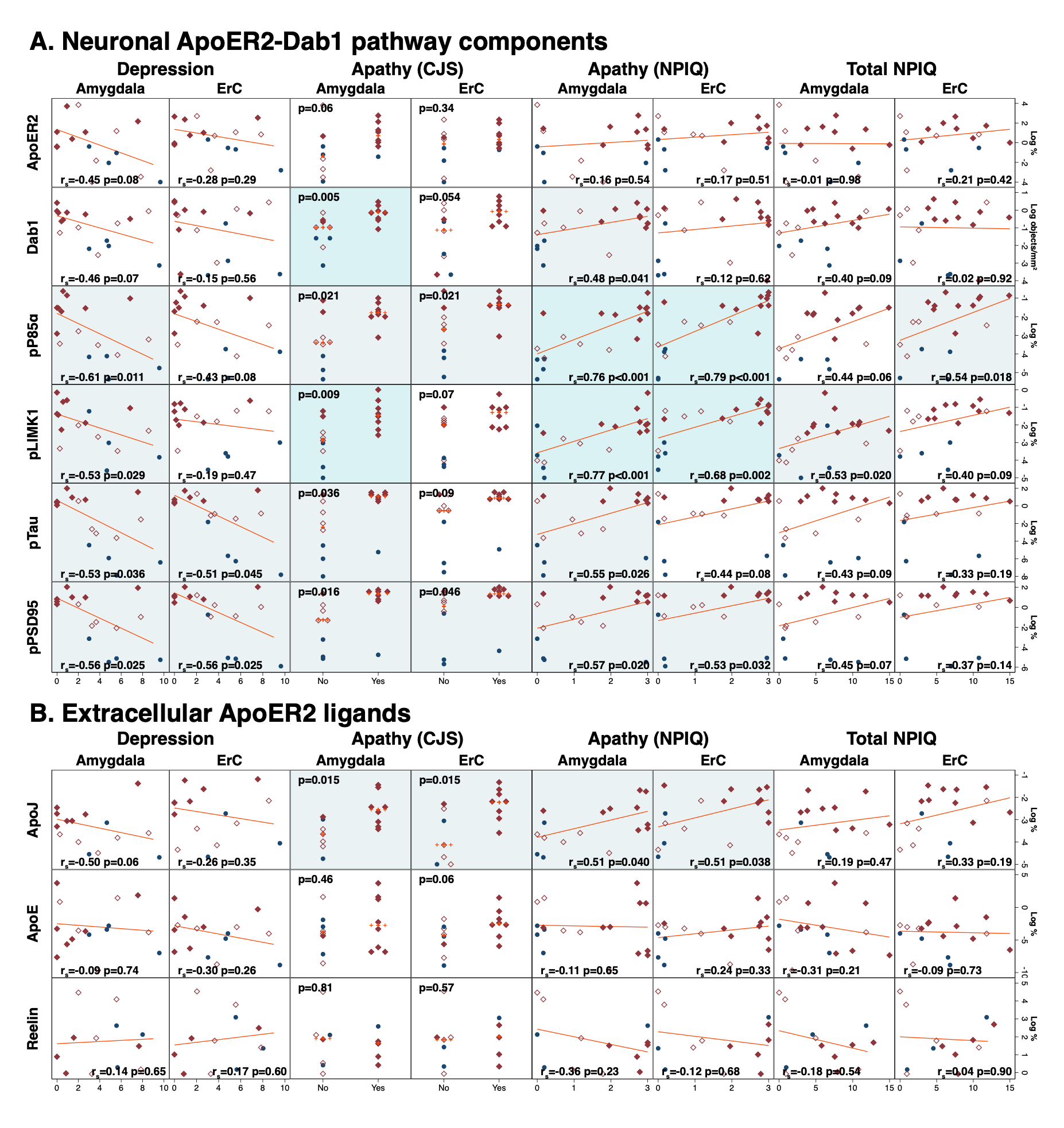

**Extended Fig 7. Correlations between ApoER2-Dab1 pathway components with depression, apathy and total NPI-Q**

In amygdala, the expression levels of five neuronal ApoER2 signaling partners **(A)** and one extracellular ApoER2 ligand (ApoJ) **(B)** positively correlated with apathy as measured by the CJS or NPIQ. In ErC, expression levels of several ApoER2-Dab1 pathway components correlated with apathy; however, these associations tended to be weaker and less robust. Dark, light blue, and white background indicate p<0.01, p<0.05, and p>0.05, respectively.

**III. Supplementary Materials**

**and Methods**

**Neuropathological Assessments**

The neuropathological assessments and endpoints captured by BBDP are detailed in a previous publication [6]. Briefly, the Braak neurofibrillary stage (0 - VI) was determined using thick 40 – 80-micron sections stained with Gallyas, Campbell-Switzer and thioflavin S stains as originally defined by Braak and Braak [7]. Senile plaque density—including neuritic, cored, and diffuse plaques—was assessed in standard regions of the frontal, temporal, and parietal lobes, hippocampal CA1 region and entorhinal/transentorhinal region. Each region was assigned a semi-quantitative score of none, sparse, moderate and frequent and converted to numerical values 0 – 3, according to the CERAD templates [1]. Plaque total is the arithmetic sum of scores from these five regions ranged from 0 – 15. Neurofibrillary tangle density was assessed in the same five regions, with CERAD templates used to obtain semi-quantitative scores of none, sparse, moderate and frequent and these are converted to numerical values 0 – 3. Tangle total is the arithmetic sum of scores from these five regions ranged from 0 – 15. The NIA-Reagan [2] consensus recommendations were used for postmortem diagnosis of AD with high, intermediate and low referring to the likelihood that dementia, if present, is due to AD histopathology. AD was at a minimum defined as intermediate or high NIA-Reagan criteria. Mild Cognitive Impairment denoted the presence of this diagnosis at the time of death. A control designation is a participant without dementia or parkinsonism during life and without a major neuropathological diagnosis.

*Braak Stage:* Describes topographical progression of neurofibrillary tangles, dystrophic neurites and neuropil threads, throughout transentorhinal and entorhinal areas, CA1 subfield of hippocampus, amygdala and cerebral neocortex. Evaluations were made, similarly as the original publication [7], in large (3 cm x 5 cm) thick (40 or 80 µm) sections stained with the Campbell-Switzer silver stain, Gallyas silver stain and Thioflavin S stains. Final judgment of tangle density is made on the basis of combined impression from all three stains. For three years, all cases were also stained with the AT8 antibody for phosphorylated tau protein. Note that the AT8 stain has been reported to give higher Braak stages as more neurites are apparent [8].

*Tangle Total:* Average neurofibrillary tangle density in the cortex of the frontal lobe, including superior, middle, and inferior frontal gyri; cortex of the temporal lobe; cortex of the parietal lobe; CA1 subfield of hippocampus; and entorhinal cortex. Tangle density scored according to the CERAD templates [1], as described for the Braak stage above.

*Plaque Total:* Average senile (amyloid) plaque density (all types of plaques considered together) in the cortex of frontal lobe, including superior, middle, and inferior frontal gyri; cortex of temporal lobe; cortex of parietal lobe; CA1 subfield of hippocampus; and entorhinal cortex. Plaque density scored according to CERAD templates [1], using large (3 cm x 5 cm) thick (40 or 80 µm) sections stained with Campbell-Switzer and Gallyas silver stains, and Thioflavin S stains. Validity and accuracy of this combination for estimating density of Aβ deposits established in BBDP laboratories through strong correlations with autoradiographic binding of Florbetapir (amyloid imaging ligand), with biochemical measures (ELISA) of Aβ in human cerebral cortex extracts and with quantitative measures (percentage of section area occupied) of an immunohistochemical stain for Aβ [6].

*Neuritic Plaque Density:* Greatest neuritic plaque density observed across frontal, temporal and parietal cortex regions, scored according to CERAD templates [1]. Evaluations made in large (3 cm x 5 cm) thick (40 or 80 µm) sections stained with the Campbell-Switzer silver stain, Gallyas silver stain and Thioflavin S stains. Final judgment of plaque density made on the basis of the combined impression from all three stains [6].

**Cognitive Assessments**

The cognitive examinations and endpoints captured by BBDP are detailed in a previous publication [6] and are summarized below.

*MMSE Test Score:* Folstein Mini Mental State Examination score (0-30) obtained most proximal to death; includes MMSE scores obtained through BBDP research clinical visits and by review of private medical records [9].

*CDR Sum:* Sum of Boxes (subsection) of the Clinical Dementia Rating (CDR) Scale. CDR is widely used for staging dementia severity [10].

*FAST Score:* FAST is a functional assessment based both on caregiver report and clinician's observations: 1 = normal; 2 = subjective (only) report of forgetfulness or work difficulties; 3 = observed early executive dysfunction; 4 = definite memory and/or executive dysfunction; 5 = some decline in basic activities of daily living (ADLs); 6 (a-e) definite decline in basic ADLs, with the final stage (e) being fecal incontinence; 7 (a-e) progressive loss of speech and motor abilities, with the final stage (e) being loss of the ability to hold up the head independently [11].

*Figure Recall Score:* Subject is asked to copy three simple figures, and after delay, is asked to reproduce the figures from memory. The score is the number correctly reproduced.

*Rey Auditory Verbal Learning Test (AVLT):* Evaluates short-term auditory-verbal memory, rate of learning, learning strategies, retroactive, and proactive interference, presence of confabulation, of confusion in memory processes, retention of information, and differences between learning and retrieval. Participants are given list of 15 unrelated words repeated over five trials and asked to repeat as many words as possible. After the five trials, the number of recalled words is summed as "AVLT Total Learning Score" (0-75 scale). After the fifth learning trial, another list of 15 unrelated words is provided. The participant recalls as many words as possible from this distracter list. After a brief delay, the participant is asked to repeat the original list of 15 words (AVLT A6) [12, 13].
